# Personalized chronic adaptive deep brain stimulation outperforms conventional stimulation in Parkinson’s disease

**DOI:** 10.1101/2023.08.03.23293450

**Authors:** Carina R Oehrn, Stephanie Cernera, Lauren H Hammer, Maria Shcherbakova, Jiaang Yao, Amelia Hahn, Sarah Wang, Jill L Ostrem, Simon Little, Philip A Starr

## Abstract

Deep brain stimulation is a widely used therapy for Parkinson’s disease (PD) but currently lacks dynamic responsiveness to changing clinical and neural states. Feedback control has the potential to improve therapeutic effectiveness, but optimal control strategy and additional benefits of “adaptive” neurostimulation are unclear. We implemented adaptive subthalamic nucleus stimulation, controlled by subthalamic or cortical signals, in three PD patients (five hemispheres) during normal daily life. We identified neurophysiological biomarkers of residual motor fluctuations using data-driven analyses of field potentials over a wide frequency range and varying stimulation amplitudes. Narrowband gamma oscillations (65-70 Hz) at either site emerged as the best control signal for sensing during stimulation. A blinded, randomized trial demonstrated improved motor symptoms and quality of life compared to clinically optimized standard stimulation. Our approach highlights the promise of personalized adaptive neurostimulation based on data-driven selection of control signals and may be applied to other neurological disorders.

## 2. Introduction

Deep brain stimulation (DBS) is a standard therapy for advanced movement disorders and is under investigation for several neuropsychiatric conditions^1^. Conventional DBS therapy is delivered with constant stimulation parameters (cDBS), unresponsive to patient activities or to variations in severity of symptoms during daily life. Thus, there is significant interest in adaptive DBS (aDBS) that uses real-time detection of neural signals to automatically adjust stimulation amplitude or other parameters in response to patients’ needs^2, 3^. Fully implantable bidirectional neural interfaces, which can sense neural activity during stimulation and have circuitry to implement feedback control, have recently become available for investigational^4, 5^ and commercial^6^ use. This development has catalyzed work in chronic invasive brain sensing^7^ and now offers the technical capability to provide chronic adaptive neurostimulation^8, 9^. However, several barriers have impeded its implementation^10^, including the limited understanding of neural signatures of specific symptoms in brain disorders treatable by DBS, technical complexity of sensing brain signals during ongoing electrical stimulation^11^, and lack of standardized algorithms for optimizing feedback control in the setting of a large parameter space.

Parkinson’s disease (PD) is a highly prevalent neurodegenerative disease and affects ∼1% of people aged 60 years or older in high-income countries^12^. Stimulation of the subthalamic nucleus (STN) via cDBS is widely used and supported by extensive class I evidence^13–16^. However, even after optimization of stimulation parameters by an expert clinician, cDBS can be associated with periods of under- and over-stimulation reflected in residual fluctuations between hypo- and hyperkinetic motor signs, such as bradykinesia and dyskinesia. This suggests individuals with PD could further benefit from aDBS. STN local field potential oscillations in a predefined beta band (13-30 Hz) are often proposed for adaptive (closed-loop) control in PD^2, 17, 18^ based on the observation that resting subthalamic beta activity is elevated in the rigid/akinetic state and reduced when motor signs are alleviated by dopaminergic medication^19^ or neurostimulation^20^. However, previous studies identified STN beta oscillations as control signals in the absence of stimulation. Motor cortical signals have also shown promise in encoding motor state and controlling aDBS^9, 21, 22^, yet the effect of stimulation amplitude on the proposed control signals was not systematically assessed. Since electrical stimulation profoundly alters oscillatory activity in the motor network^20, 23^, it is crucial to define neural biomarkers that can still be measured and tracked during stimulation at therapeutic amplitudes, a scenario which is to date under-explored.

Brief studies of invasive neurophysiological control signals, often using externalized brain leads or distributed control through external computers, have demonstrated that aDBS in PD can match or exceed the benefit provided by cDBS^24–26^. However, these results were derived from in-laboratory studies and group-level analyses, lacking individualized, data-driven approaches. It remains unclear whether aDBS provides improvement beyond that of clinically optimized cDBS in real-life naturalistic settings. Here, we developed a data-driven analysis pipeline to identify individualized neural biomarkers of PD symptoms and engineered personalized aDBS algorithms that did not pre-select STN beta or other frequency bands. Our pipeline identified finely-tuned gamma oscillations (65-70 Hz)^27^ either in STN or sensorimotor cortex as the optimal biomarker of residual fluctuations in motor function that were still robust during varying stimulation amplitudes. In a blinded, randomized study across one month per condition, we demonstrate for the first time that adaptive stimulation reduces the time spent with bothersome motor signs compared to clinically optimized continuous stimulation.

## 3. Results

We recruited three patients with PD from a population undergoing DBS implantation for motor fluctuations. All patients underwent bilateral placement of quadripolar DBS leads into the STN and quadripolar paddles into the subdural space over the sensorimotor cortex (Fig. 1a-b, Extended Data Fig. 1). Cortical recordings were performed using non-overlapping bipolar pairs with the anterior montage (montage 1) having at least one electrode covering the precentral gyrus and the posterior montage (montage 2) having at least one electrode on the postcentral gyrus (Extended Data Fig. 1a, c-g). Leads were connected to an investigational bidirectional neural interface (Medtronic Summit RC+S). This device is capable of chronically streaming high-resolution time domain data in naturalistic settings while providing therapeutic stimulation and can perform aDBS using fully embedded algorithms^5, 9^. We developed adaptive algorithms tailored to the specific clinical needs of each patient. Before patients began the aDBS trial, standard of care cDBS was optimized by a movement disorder specialist over a range of 11-31 months (mean±standard deviation 22±10), including at-home self-optimization by the patients. Patients identified their most bothersome motor symptom persisting on clinically optimized cDBS, e.g., bradykinesia (Fig. 1c; Fig. 2). In addition, we defined the most prominent symptom in the opposite motor state (e.g., the hyperkinetic symptom dyskinesia if the hypokinetic symptom bradykinesia was the most bothersome motor sign). This was done to ensure that aDBS did not improve hypokinetic symptoms at the expense of hyperkinetic symptoms or vice versa. For one patient, bothersome residual motor fluctuations were restricted to one side of the body (pat-1). The remaining two patients perceived persisting bilateral symptoms (pat-2 and pat-3). To identify the optimal stimulation limits for symptom control during aDBS treatment, we determined the high and low stimulation amplitudes needed to address the patients’ hypo- and hyperkinetic states, respectively (Fig. 2). As stimulation impacts neural activity within the stimulated networks, we identified neural biomarkers during active stimulation.

**Figure 1.**
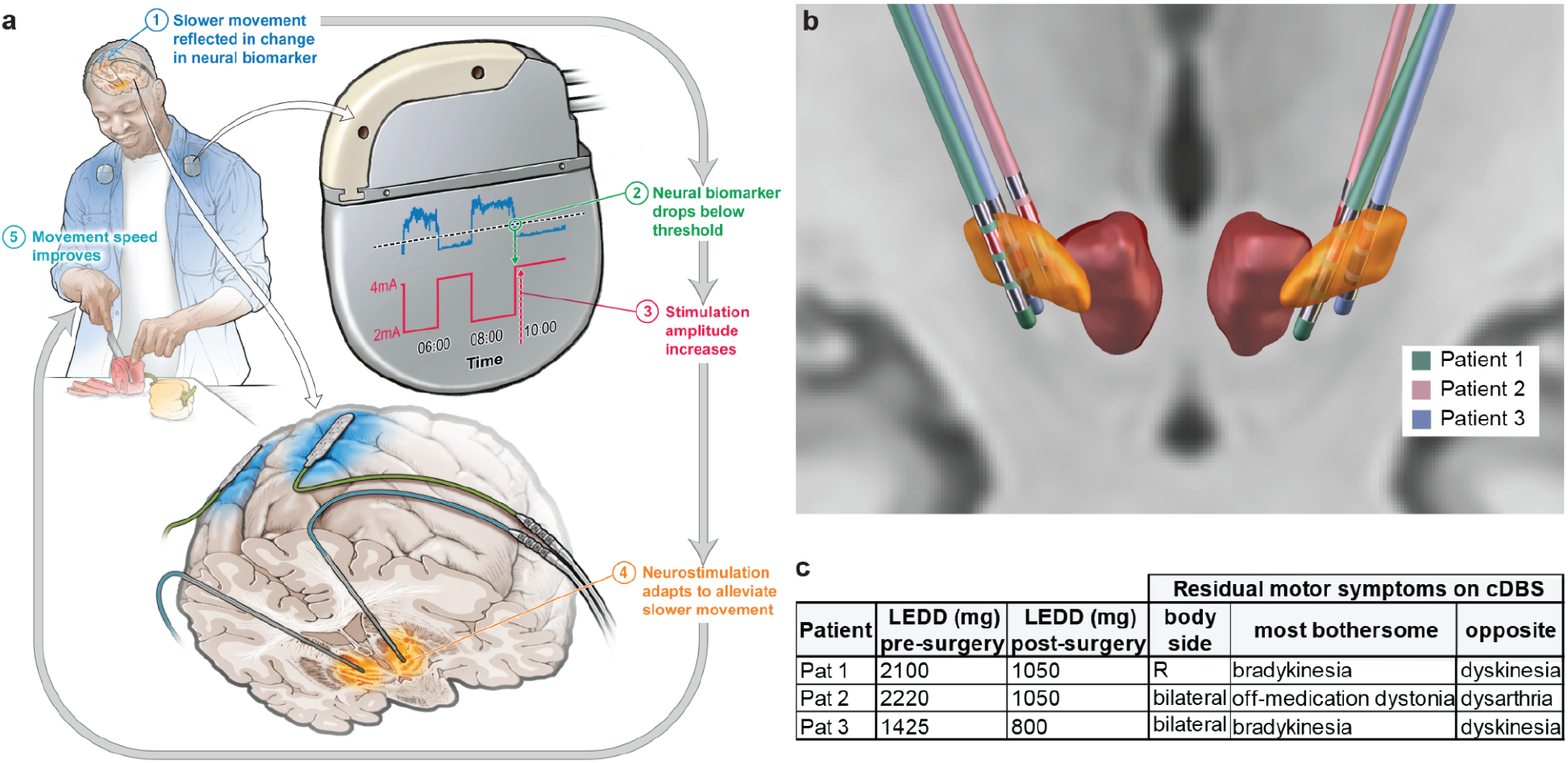
Configuration of implanted hardware, algorithmic model and patient demographics. **a**, Illustration of the adaptive paradigm starting with real-life sensing of brain activity (blue) that reflects changes in patient’s mobility–in this example slowness of movement (bradykinesia). Neural activity is sensed continuously on-board the DBS device from either the STN or sensorimotor cortex using depth or subdural electrodes, respectively. Here, we illustrate an example of a cortical control signal for fully-embedded adaptive implementation. Once a change in the brain signal across a predefined threshold is detected (green), the stimulation amplitude increases or decreases automatically (red) at the target brain region (STN). This adaptation of stimulation amplitude to the patient’s needs leads to improved symptoms–in this example, increased stimulation amplitude results in faster movement. **b**, Localization of depth leads in the STN with active contacts colored in red across patients in normalized Montreal Neurological Institute space. STN is highlighted in orange and the red nucleus in red. **c**, Patient characteristics including pre- and post-surgery levodopa equivalent daily dose (LEDD, mg) and residual motor fluctuations on clinically optimized cDBS, including the body side, the most bothersome symptom and the “opposite” symptom, referring to the opposite medication state, such as hyperkinetic symptoms for hypokinetic bothersome symptoms, or effects of DBS that limit the therapeutic window (e.g., effects on speech).

**Figure 2.**
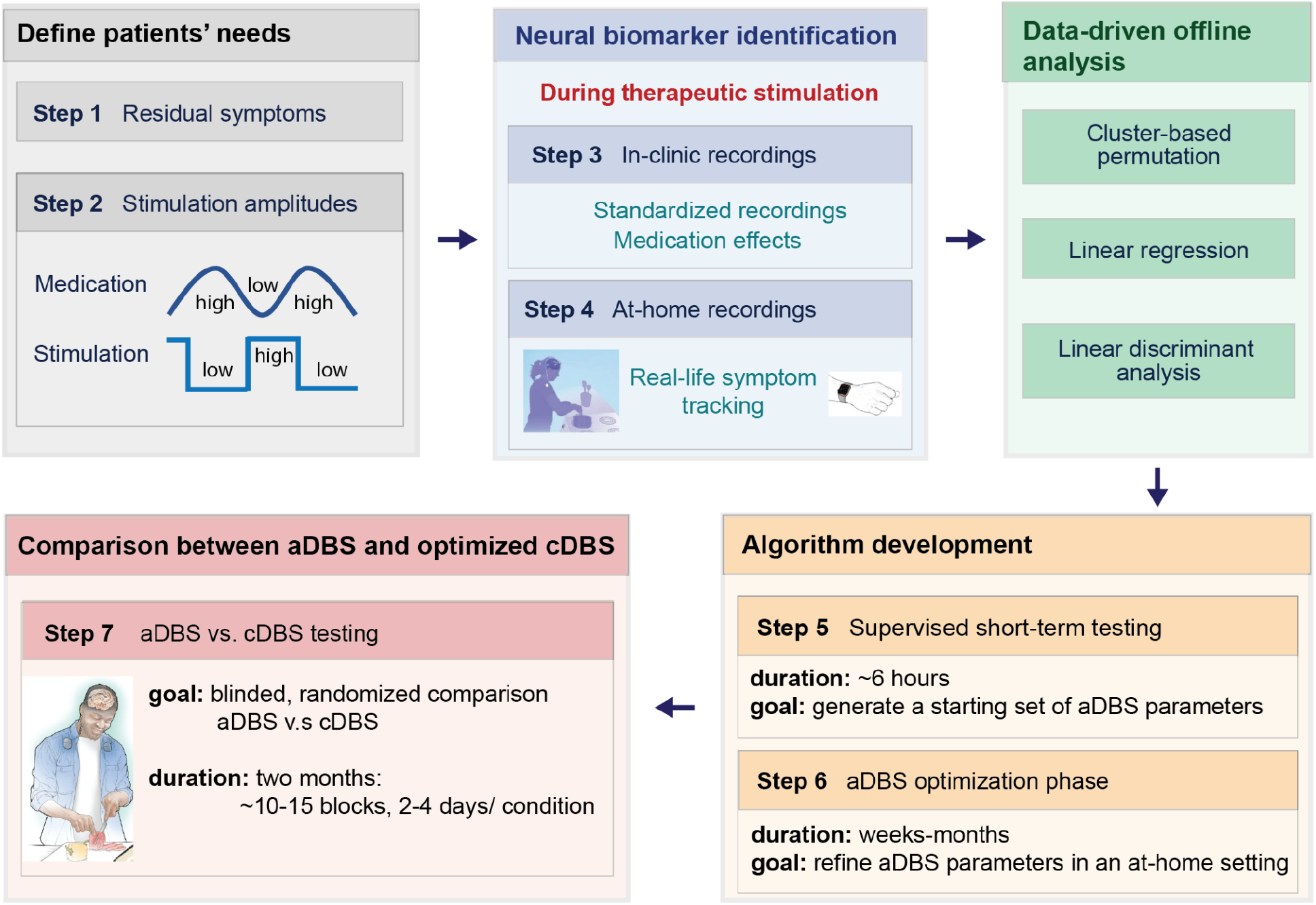
Workflow for data-driven biomarker identification and aDBS implementation. We employed a workflow consisting of seven steps. These steps involved identifying bothersome symptoms and required stimulation amplitudes for symptom control for each patient (steps 1 and 2), in-clinic and at-home neural recordings with simultaneous symptom monitoring for biomarker identification (steps 3-4) and refining parameters for patient-tailored adaptive algorithms using supervised short-term (step 5) and long-term (step 6) at-home testing. The workflow culminated in blinded, randomized comparisons between cDBS and aDBS in multiple blocks of 2-4 days per condition (total of one month per condition) in patients’ real-life environments (step 7).

### Motor network gamma oscillations identified as optimal control signals

We then developed a data-driven pipeline to identify electrophysiological field potentials that correlated with patient symptoms and implemented them as feedback signals for the adaptive algorithms. Our workflow consisted of seven formalized steps that included streaming neural data in-clinic and at-home during active stimulation, leading to a blinded, randomized comparison between cDBS and aDBS for several weeks in patients’ naturalistic environments (Fig. 2). We utilized a combination of non-parametric statistics and machine learning methods to search the frequency space in both STN and sensorimotor cortex for the physiological signals that optimally predicted the occurrence of patients’ most bothersome motor signs (see methods). Converging evidence from in-clinic and at-home recordings demonstrated that finely-tuned gamma (FTG) oscillations performed optimally as a predictor of medication related symptom state^21^. In the setting of subthalamic stimulation above a certain amplitude, the frequency of FTG oscillations within their typical 60-90 Hz range often shifts to a subharmonic of stimulation frequency, behaving as a “driven oscillator” (Fig. 3a)^28^. Despite the potential susceptibility to artifacts when sensing neural signals during active stimulation, we demonstrated that FTG oscillations at half the stimulation frequency were not artifactual, as they represented the entrainment of levodopa-induced narrowband gamma oscillations that were observed off stimulation (Fig. 3a)^21^.

**Figure 3.**
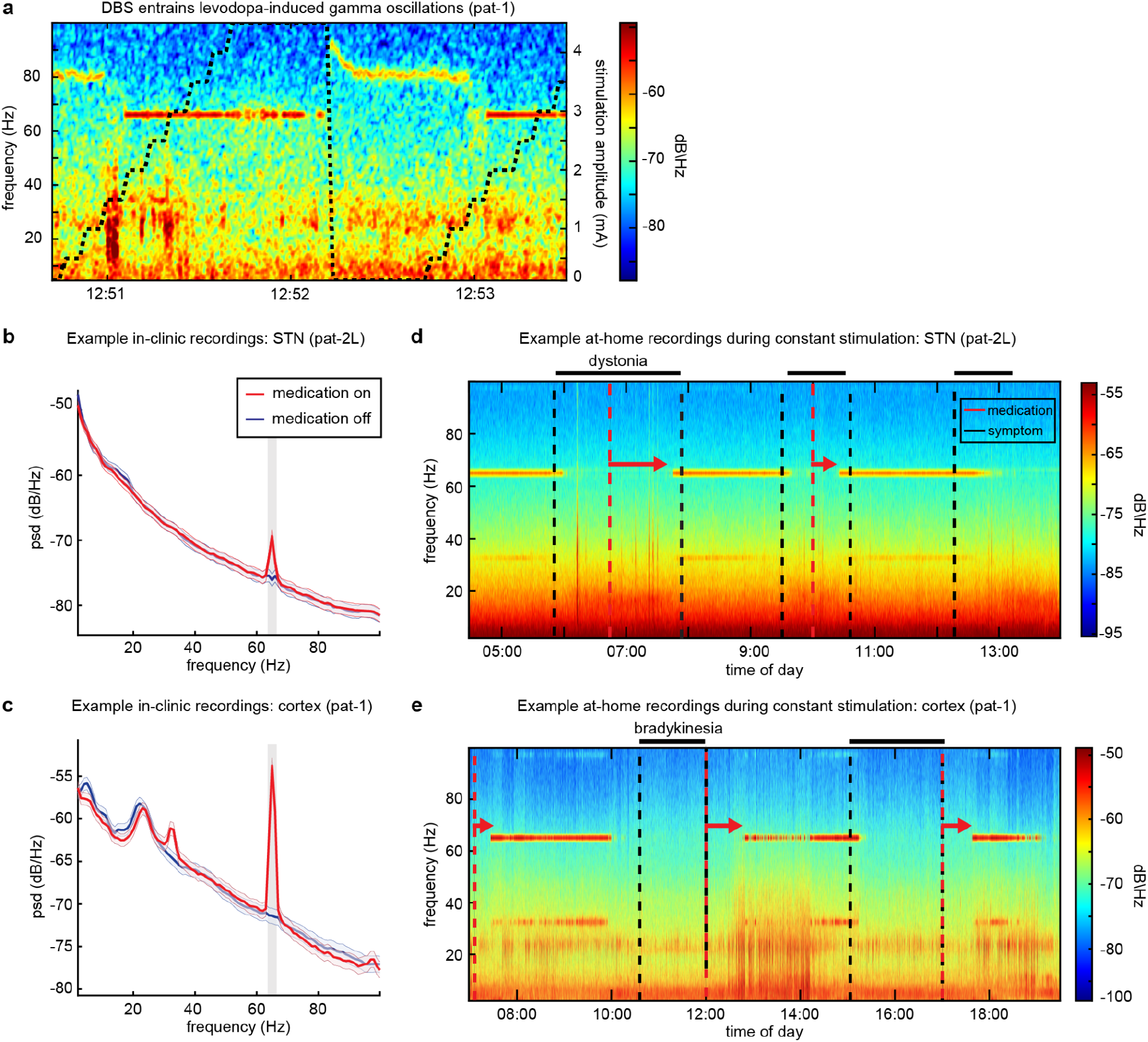
Examples of stimulation-entrained finely-tuned gamma oscillations in both in-clinic and at-home recordings. **a**, Example spectrogram of cortical activity in the on-medication state during systematic variations in stimulation amplitude (black dotted line), illustrating the phenomenon of stimulation-induced entrainment of gamma oscillations at half of the stimulation frequency (pat-1). Levodopa-induced finely-tuned gamma (FTG) oscillations occur at 80-90 Hz when stimulation amplitudes are low but become entrained to half the stimulation frequency (65 Hz) when stimulation exceeds a certain amplitude (1.5 mA in this example). **b-c**, Examples of biomarker identification using standardized in-clinic neural recordings (**b,** pat-2L, **c,** pat-1). Plots show power spectra during on- and off-levodopa states (mean±standard error of the mean), i.e., periods during which hyper- and hypokinetic symptoms would emerge, respectively. Recordings are collapsed across low and high stimulation amplitude conditions, which were both amplitudes at which FTG entrained to half the stimulation frequency. We found that medication yielded the largest effect on entrained finely-tuned gamma power at half the stimulation frequency in the STN (**b**, pat-2L) and motor cortex (**c**, pat-1) when controlling for effects of stimulation (Extended Data Fig. 2). Significant clusters are highlighted in gray. **d-e**, At-home recordings during constant stimulation amplitude and patients’ normal medication schedule in the STN (**d**, pat-2L) and motor cortex (**e**, pat-1). Patients marked their medication intake (red dashed line) and on- and off-set of their most bothersome symptom in their motor diary (completed in 30-minute intervals) and the streaming application^9^. Both patients had bothersome off-state symptoms, lower limb-dystonia (**d**, pat-2) and bradykinesia (**e**, pat-1). For both, FTG oscillations occur ∼45 minutes after medication intake (red arrows), corresponding to a typical latency of onset for dopaminergic medication. When the patient marked their most bothersome symptom (indicated by the black dashed line) in their motor diary, FTG oscillations disappeared, indicating a transition to an off-medication state.

FTG oscillations were prominent during hyperkinetic states (Fig. 3-4, Extended Data Fig. 2a, Extended Data Fig. 3a-b), were reduced by 33-96% (median±standard deviation 90±28%) when these states ended, and could track symptom changes over the full range of patient-specific stimulation amplitudes to be applied during aDBS (Extended Data Fig. 2a, Extended Data Fig. 3a-b). Similar to levodopa-induced FTG, entrained gamma power fluctuated with medication state in-clinic (Fig. 3b-c, Fig. 4a-b, Extended Data Fig. 2a) and at-home while stimulation remained unchanged (Fig. 3d-e), and predicted hyperkinetic symptoms (Fig. 4c-d, Extended Data Fig. 3a-b). Subthalamic beta activity is often proposed as an optimal control signal for aDBS, and consistent with the literature, we observed beta power peaks off stimulation and off medication in four of five hemispheres (Extended Data Fig. 2c). However, medication effects diminished with active stimulation (Extended Data Fig. 2d) and were only significant in one patient (Fig. 4a, Extended Data Fig. 2b). Further, beta power did not reliably track symptom fluctuations during chronic stimulation in the home environment with the exception of one hemisphere (Fig. 4c-d, Extended Data Fig. 3c-d). Using STN beta power in conjunction with subthalamic or cortical FTG resulted in only a minimal change in symptom prediction accuracy (Fig. 4e, AUC increased from 0.75±0.07 to 0.76±0.07, mean across subjects ± standard deviation) and we were not able to show a significant difference between adding STN beta versus alternative random frequency bands for any subject (range of *p*-values: 0.055-0.75).

**Figure 4.**
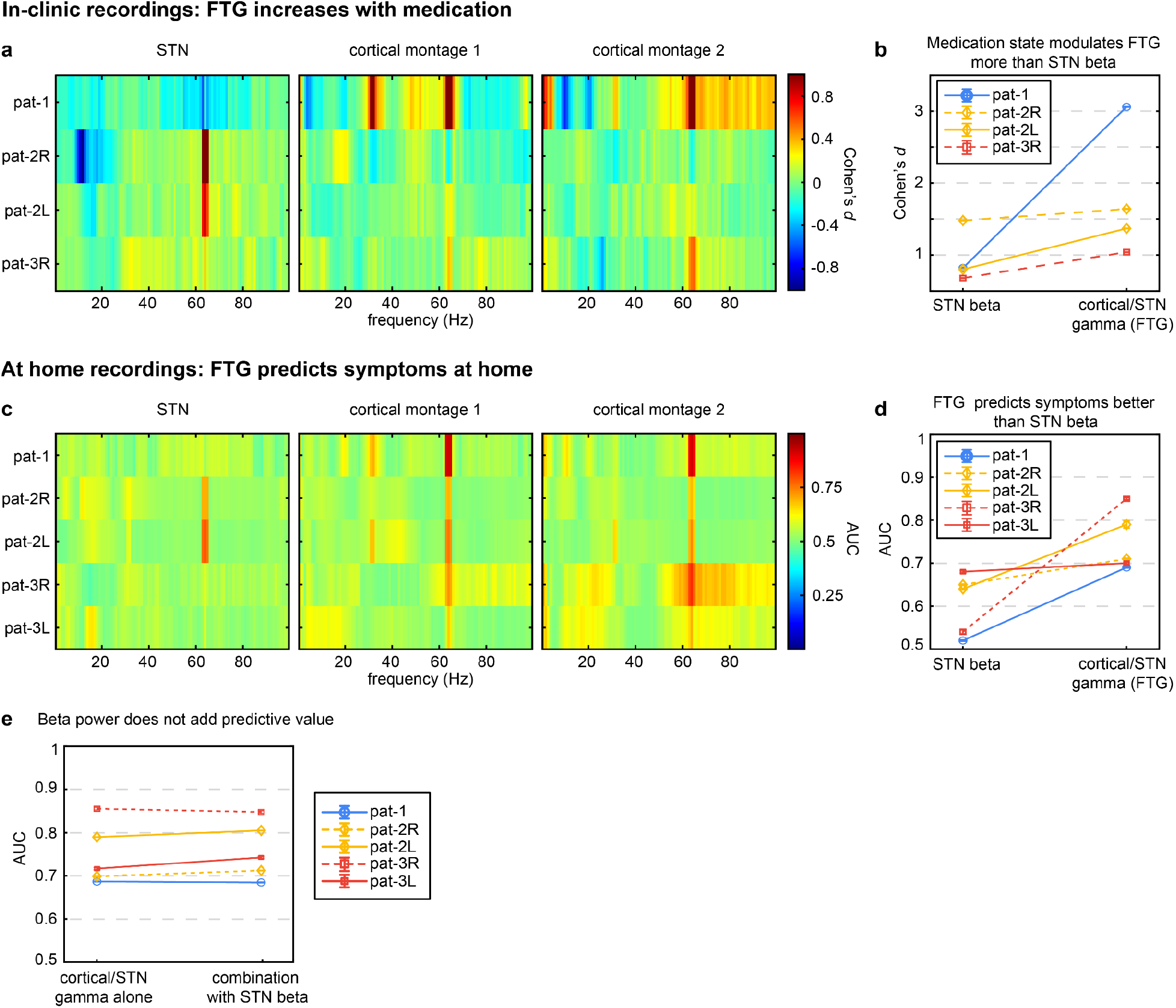
Data-driven biomarker identification for all hemispheres. **a-b**, Results of the within-subject nonparametric cluster-based permutation analysis for in-clinic recordings. **a**, Graphs illustrate the effect size (Cohen’s *d*) of the main effect of medication on power as a function of frequency in the STN (left), cortical montage 1 (middle), and cortical montage 2 (right) for all patients. Red and blue colors represent positive effects (medication on > medication off) and negative effects (medication off > medication on), respectively. For all patients, we found finely-tuned gamma (FTG) oscillations in the STN (pat-2, both hemispheres) and cortex (pat-1, left hemisphere and pat-3, right hemisphere) to be the optimal biomarker for medication-related fluctuations during active stimulation (Extended Data Fig. 2). We did not find any significant effects in the left hemisphere of pat-3. **b**, The effect sizes for cortical and STN FTG oscillations (right) were superior to those for STN beta oscillations (left) for all patients (mean±standard error of the mean across permutations). **c-e**, Results of the within-subject linear discriminant analysis for at-home recordings using power spectral density at the three brain sites to predict the occurrence of the most bothersome symptom. **c**, The three graphs illustrate the initial area under the curve (AUC) prior to bandwidth optimization as a function of frequency for the STN (left), cortical montage 1 (middle), and cortical montage 2 (right) for all patients. Across patients, we show that FTG oscillations in the STN (pat-2, both hemispheres) and cortex (pat-1, left hemisphere and pat-3, both hemispheres) were the best predictors of the occurrence of the most bothersome symptom and superior to beta oscillations (**d**, mean±standard error of the mean across permutations, Extended Data Fig. 3). **e**, The combined use of STN/cortical gamma and STN beta bands provided minimal improvement in the AUC of at-home symptom prediction using linear discriminant analysis (mean±standard error of the mean across permutations).

### Adaptive stimulation algorithm tracked residual motor fluctuations

During aDBS, we used STN or cortical FTG at half stimulation frequency as the control signal for all patients (Fig. 5a). FTG oscillations represented hyperkinetic states; therefore, we designed aDBS algorithms that reduced stimulation amplitude when the control signal was high to either avoid hyperkinetic symptoms such as dyskinesia (pat-1 and pat-3) or relieve stimulation-induced side effects such as dysarthria (pat-2) (Fig. 5b-c). In total, patients spent 74.9±12.1% of their awake time (mean±standard deviation) in the high amplitude state, nearly three times longer than in the low amplitude state (25.1±12.2%, Fig. 5d). This is congruent with patients’ self-reported symptoms during blinded at-home cDBS testing, where time spent with hypokinetic symptoms was on average 3.7 times longer than with hyperkinetic symptoms. Our algorithms acted on a timescale of minutes to hours, consistent with established carbidopa-levodopa pharmacokinetics^29^. On average, high amplitude stimulation states (responding to hypokinetic clinical states) lasted 1.5±0.27 consecutive hours and low amplitude stimulation states (responding to hyperkinetic symptoms, Fig. 5e) lasted 0.65±0.48 hours. Patient 3 received recurring short epochs of low stimulation amplitudes on aDBS due to brief but frequent bouts of dyskinesia on cDBS (Fig. 5d,e). Across waking hours, all patients received greater total electrical energy delivered (TEED) while on aDBS compared to cDBS (15.1±20.4% increase from cDBS; Fig. 5f). The majority of nighttime was spent at the high stimulation amplitude (96.2±1.4%), which reflected suppression of FTG during sleep and thereby resulted in greater nighttime TEED compared to cDBS (35.5±33.9% increase from cDBS; Extended Data Fig. 7a-b). In contrast to our biomarker for hyperkinetic states (i.e., FTG), use of a biomarker representing hypokinetic states (e.g., STN beta oscillations), for which stimulation amplitude is high when the biomarker is high, would likely result in lower stimulation amplitude during sleep since it is suppressed.

**Figure 5.**
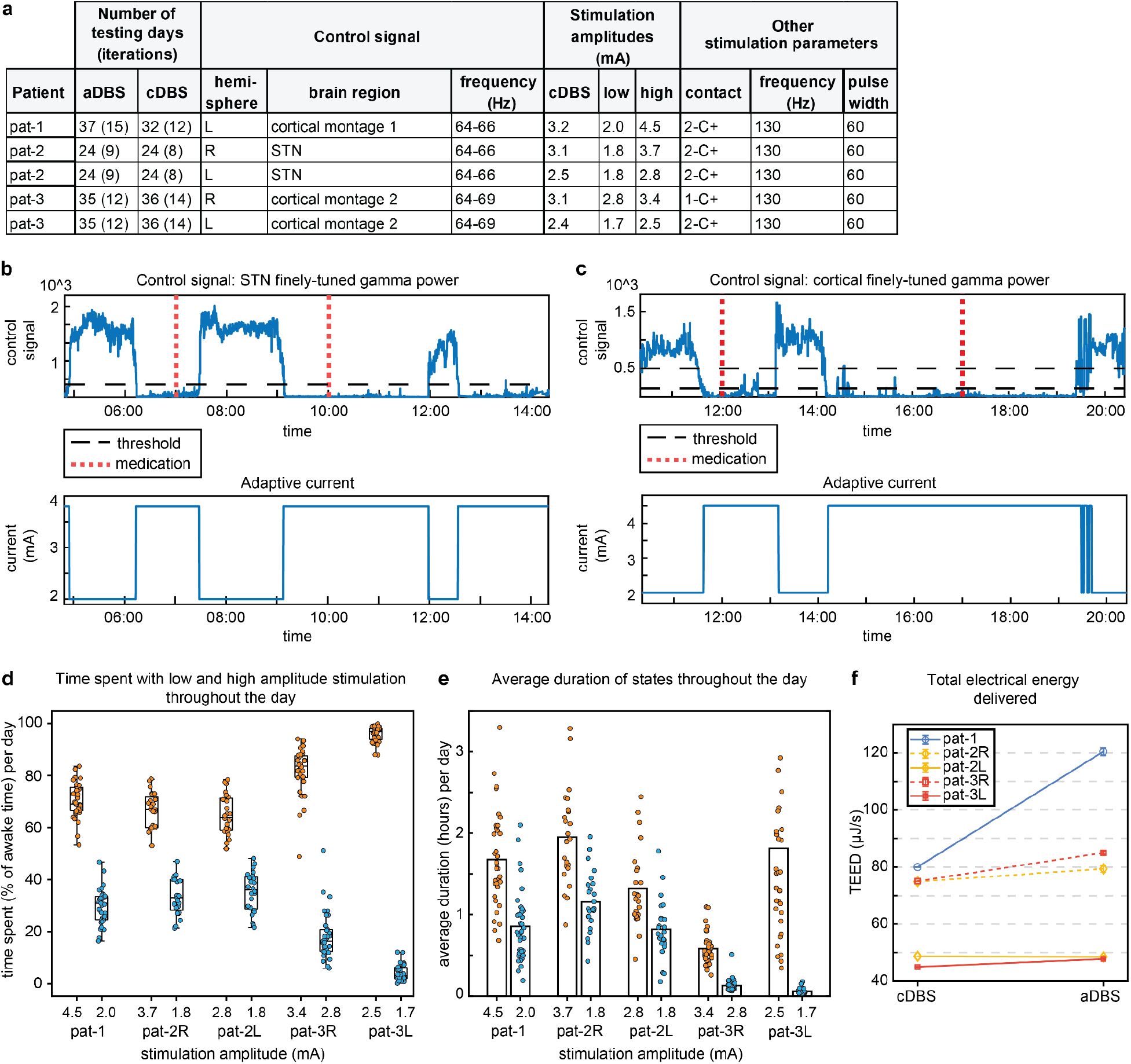
Characteristics and technical performance of adaptive DBS algorithms. **a**, Summary of the final stimulation parameters used for blinded, randomized comparisons between stimulation conditions including the control signal for aDBS. All parameters but stimulation amplitude were identical between cDBS and aDBS. **b-c**, Examples of two control algorithms using subthalamic (**b**, pat-2R) and cortical (**c**, pat-1) finely-tuned gamma oscillations at half the stimulation frequency as control signals. In each graph, the upper subplot illustrates the control signal as a function of time with thresholds (black) that are used to determine changes in stimulation amplitude. The lower subpanel illustrates the stimulation amplitudes responding to fluctuations in the neural signal. In all patients, we used an on-state biomarker, such that stimulation amplitude *decreases* when the biomarker amplitude *exceeds* a threshold. Timing of dopaminergic medication intake is marked by dashed red vertical lines. **d**,**e**, Dynamics of algorithm performance showing adaptive changes on a time course of minutes-hours. **d**, Daily percent time spent at each stimulation amplitude. **e**, Average duration of each stimulation amplitude state in a day. Each dot represents one day of aDBS testing. Pat-3’s left hemisphere’s state in the high amplitude has three outliers not currently plotted which include 4.32, 5.15, and 7.32 hours. **f**, Mean (± standard error of the mean) total electrical energy delivered (TEED) during aDBS and cDBS, showing increased TEED throughout aDBS in all patients during the day.

### Adaptive stimulation improved motor symptoms that persisted on clinically optimized continuous stimulation

We compared aDBS to clinically optimized cDBS for a cumulative period of one month per condition. Stimulation conditions were applied blindly in randomized brief blocks of several days and assessed using patient ratings of daily symptoms via digitalized questionnaire. We further tracked motor fluctuations with validated wearables^30^. We found aDBS reduced the time spent with bothersome motor symptoms compared to optimized cDBS in all three patients (Fig. 6a, each patient: *p*<0.001). Further, this improvement did not occur at the expense of the opposite symptom, which was unaffected or improved (Fig. 6b, pat-1: *p*=0.56, pat-2: *p*=1, pat-3, *p*=0.02). Patients also self-assessed symptom severity. Symptoms were less or equally severe during aDBS (Extended Data Fig. 8b-c, bothersome symptoms: pat-1: *p*<0.001, pat-2: *p*=0.18, pat-3: *p*=0.93; opposite symptoms: pat-1: *p*=0.18, pat-2: *p*=1, pat-3: *p*=0.93). For the two patients with upper limb symptoms, for whom bothersome symptoms could be tracked with wrist-watch style wearable monitors, objective metrics confirmed the reduction in motor fluctuations (Fig. 6d-e). Adaptive DBS decreased the degree of motor fluctuations throughout the day as measured by the difference between symptom severity during hypo-versus hyperkinetic states (bradykinesia: pat-1: *p*<0.001, pat-3 left body: *p*=0.005, pat-3 right body: *p*=0.046; dyskinesia: pat-1: *p*=0.04, pat-3 right body: *p*=0.03). This was a result of a significant reduction of bradykinesia severity during hypokinetic states (pat-1: *p*=0.004) and dyskinesia severity during hyperkinetic states (pat-3 right body: *p*=0.04).

**Figure 6.**
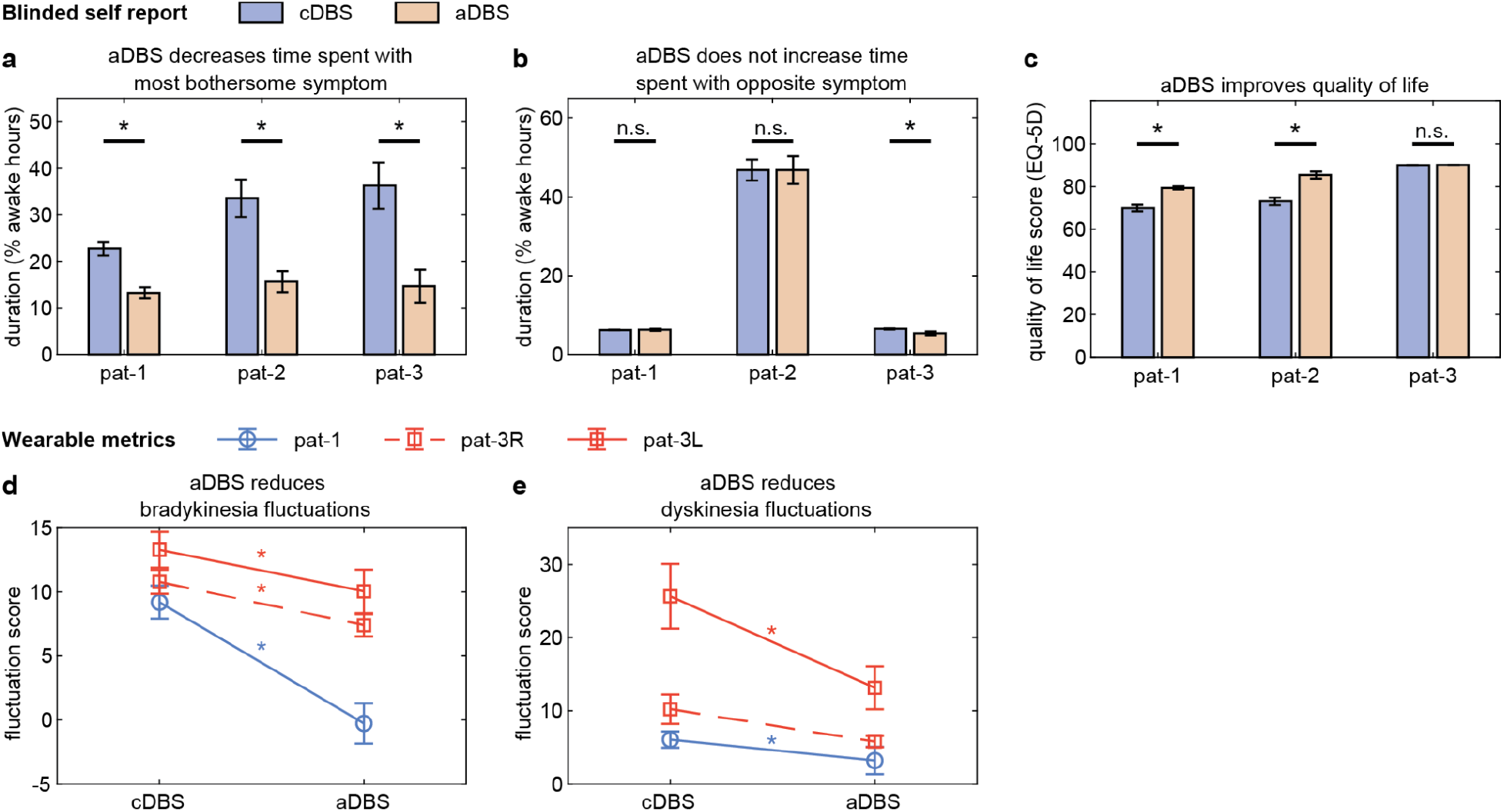
Effects of aDBS compared to cDBS on both subjective and objective metrics of motor symptoms and quality of life. **a-b**, Self-reported symptom duration from daily questionnaires. **a**, aDBS resulted in a significantly decreased percentage of awake hours experiencing the most bothersome symptom (each patient *p*<0.001). **b**, Even while reducing time with the most bothersome motor symptom, aDBS resulted in either no significant change (pat-1: *p*=0.56, pat-2: *p*=1) or an improvement (pat-3: *p*=0.02) in the percentage of awake hours experiencing the opposite symptom. **c**, Quality of life, as measured by the EQ-5D, was improved for two of three patients (pat-1 and pat-2: *p*<0.001). The third patient reported very high quality of life scores, with minimal reported variance for both cDBS and aDBS. **d-e**, Wearable monitor scores demonstrating the decreases in symptom intensity fluctuations. Only patients 1 and 3 are displayed, as patient 2’s bothersome and opposite symptoms were not measurable by a wearable device. Laterality refers to the brain hemisphere where aDBS was applied (and therefore contralateral motor sign measurement). **d**, Fluctuation scores represent differences between wearable scores during hypo- and hyperkinetic states defined by the neural signal. Fluctuations were reduced during aDBS compared to cDBS, representing a stabilized clinical profile throughout the day (pat-1: *p*<0.001, pat-3R: *p*=0.005, pat-3L: *p*=0.046). **e**, Similar improvements in the dyskinesia fluctuation score were seen in two of three hemispheres with aDBS (pat-1: *p*=0.04, pat-3R: *p*=0.03). Error bars reflect standard error of the mean.

Additionally, aDBS was associated with improved quality of life reports in two patients (Fig. 6c, pat-1: *p*<0.001, pat-2: *p*<0.001, pat-3: *p*=0.34). Quality of life metrics for patient 3 may have exhibited a ceiling effect, as they reported high baseline quality of life metrics (EQ-5D, overall health: ∼90%) and low variability despite reporting bothersome levels of residual bradykinesia on cDBS. Adaptive DBS did not adversely affect any additionally monitored motor symptoms, and instead improved gait disturbance for patient 2 (Extended Data Fig. 8d-i, pat-1 all symptoms *p*>0.08, pat-2 gait *p*<0.001, pat-2 symptoms besides gait *p*>0.11, pat-3 all symptoms *p*>0.85). Adaptive DBS also did not adversely affect any monitored non-motor symptoms (depression, anxiety, apathy, impulsivity, pat-1: *p*>0.62, pat-2: *p*=1, pat-3: *p*=1). None of the patients reported perceiving unusual sensations or changes in stimulation while on adaptive stimulation.

## 4. Discussion

We developed a data-driven pipeline for the implementation of adaptive DBS that utilized subthalamic or cortical field potentials to auto-adjust stimulation amplitudes in order to alleviate residual motor fluctuations in three individuals with Parkinson’s disease. Patients were previously clinically optimized on standard continuous DBS over several months. Our pipeline was naïve to the frequency components and detection sites of neural signal biomarkers and revealed that in all patients finely-tuned gamma oscillations performed optimally for detecting bothersome residual motor signs. In a blinded, randomized study, we demonstrate for the first time that aDBS improved motor signs and quality of life in PD during normal daily activities at home compared to standard-of-care cDBS.

### Adaptive versus contingent stimulation

In our protocol, a nonzero level of stimulation was always present, but stimulation amplitude was continuously adjusted in response to neural feedback. This form of adaptive DBS should be distinguished from “contingent” neurostimulation, where sensed neural signals can trigger a brief, pre-programmed epoch of stimulation, but sensing and stimulation are not simultaneous. Contingent neurostimulation was first introduced clinically for epilepsy^31^, is under investigational study for other neuropsychiatric disorders^32^, and is more applicable to paroxysmal disorders that do not require uninterrupted therapy. Sensing during stimulation is technically challenging, since the amplitude of the stimulus artifact is several fold higher than that of the neural signals used for adaptive control, and thus requires a different device design^33^. The successful implementation of our methodology for PD provides a framework for configuring adaptive neurostimulation that may be extended to several other neuropsychiatric disorders, many of which are currently under investigation but have not yet shown superior benefit compared to cDBS at home during normal daily activities^34^.

### Varying time scales for adaptive DBS

Adaptive neurostimulation could operate on a variety of timescales depending on the desired effects on neural circuits^35^. The original description of aDBS in PD, based on perioperative testing using externalized leads, operated on a very fast, subsecond timescale with the goal of shortening pathologically prolonged bursts of subthalamic beta activity^24, 36^. Alternatively, prolonged time scales spanning weeks to months may be appropriate in some neurological or psychiatric conditions where fluctuations in network activity underlying symptoms are slow. Here, the aDBS algorithms operated on an intermediate timescale of minutes to hours, consistent with the time course of ongoing fluctuations in Parkinson’s patients’ motor signs during cDBS (Fig. 5e).

Though DBS successfully reduces the motor fluctuations of advanced PD, patients typically still require a combination of antiparkinsonian medications (albeit reduced) and stimulation for best function^37^. Because the effective brain concentration of levodopa has peaks and valleys, under constant stimulation patients may continue to experience periods of hyperkinetic and/or hypokinetic function. While less severe than before device implantation, these residual fluctuations were bothersome in our study patients. aDBS allowed seamless integration of medication and stimulation therapy by providing less stimulation when medication is active (“on”), associated with hyperkinetic states, and more when medications wore off, i.e., during hypokinetic states.

### Optimal frequency bands for adaptive control

We identified FTG oscillations in the STN and cortex as optimal markers of residual motor signs in all three patients. FTG was first identified in the basal ganglia as a marker of the levodopa “on” state^38^, and much later found in the motor cortex in rodent models of parkinsonism^39^ and in humans^21^, where it is especially prominent during on-period dyskinesia. A remarkable aspect of FTG as an aDBS control signal is that, when stimulation is turned on, FTG peak frequency shifts slightly away from its “natural” frequency to become entrained to the nearest subharmonic of stimulation frequency, typically in a 1:2 entrainment pattern^21^ (Fig. 3a). Thus, the peak frequency is highly predictable—a useful property when choosing the frequency band to be utilized for adaptive control. While a signal appearing at an exact subharmonic of stimulation frequency may at first glance appear to be an electrical artifact, there are now multiple comprehensive arguments against an artifactual origin. Entrained FTG amplitude is strongly modulated by the physiological state of the brain, including on/off medication cycles (Fig. 3b-e) and sleep-wake cycles, is often more prominent at a site distant from the stimulating contact (cortex) than adjacent to it (STN), has a very specific topography (greater in precentral than postcentral gyrus), and, when stimulation is turned off, can require additional time to “wash out”^40^. These findings indicate a physiological origin.

Most previous in-laboratory studies of aDBS in PD employed the spectral power of subthalamic beta oscillations as a feedback signal^24, 25, 41^. The choice of STN beta band activity as a feedback signal was driven by physiology studies performed in the absence of stimulation^42^. However, in our cohort, spectral power of STN beta oscillations did not track residual motor fluctuations in the home environment in four of five hemispheres (Extended Data Fig. 3c-d). Therapeutic stimulation did reduce beta activity, similar to previous reports^20, 43, 44^, even at low-therapeutic stimulation levels (Extended Data Fig. 2d). Thus, beta band activity may no longer adequately track residual motor signs within the range of stimulation amplitudes relevant for adaptive control. A critical element of our aDBS development pipeline was to evaluate the relation of oscillatory activity to bothersome motor signs *over the full range of stimulation amplitudes to be used in adaptive control*, rather than in the off-stimulation state. In the hyperkinetic state, reliable entrainment of FTG occurred within the range of therapeutic amplitudes used during adaptive implementation, and the amplitude of entrained FTG demonstrated low variance between our defined low and high-therapeutic stimulation limits. Of note, a beta band controller may well be optimal for aDBS algorithms in PD patients designed for different time scales, such as for modification of bursts of oscillatory activity^24, 36^. Here, our subthalamic beta band detection was limited to a specific bipolar montage in which recording contacts were immediately adjacent to the active contact, whereas other bidirectional neural interface systems that allow for additional montage configurations may be more optimized for beta detection^18^.

### Cortical versus subcortical signals for adaptive DBS

Our results highlight the utility of multi-site brain recordings for aDBS algorithms. Cortical recordings did prove invaluable for two of the three patients who did not have adequate neural biomarkers in the STN. Subcortical signals may be insufficient for aDBS in certain scenarios, such as when the signal is excessively contaminated by stimulation artifacts. However, we have not proven that cortical recordings are critical for aDBS in PD. To achieve high signal-to-noise recordings, we placed cortical leads in the subdural space directly on the brain surface. This has disadvantages of adding invasiveness to the surgery and committing the patient to hardware that is not easy to remove. In future studies, cortical control signals might be obtained less invasively from electrodes placed under the scalp^45^.

### Adaptive DBS outperforms continuous DBS in real life

Most prior studies comparing cDBS and aDBS for the treatment of PD motor symptoms have been limited to highly-controlled in-clinic or laboratory settings^22, 24–26, 46^. Many were performed peri-operatively with externalized leads^24, 26, 46^, which differs greatly from the home environment with a fully implanted stimulator and decoder. Perioperative experimentation also precludes the optimization of cDBS parameters by an expert neurologist, which usually takes several months^47^, and therefore does not represent a rigorous comparison to actual standard of care. We recently described a case report suggesting benefit of home-based embedded aDBS in one patient^9^, but the study was restricted to four days in an unblinded subject.

Here, we addressed these limitations and implemented aDBS algorithms in naturalistic settings for multiple weeks during patients’ routine daily activities, including work, travel, and sleep. The algorithm was tested repeatedly in short blocks of several days to minimize systematic confounds from situational influences on patient ratings. We personalized each algorithm to the patients’ specific clinical needs and compared aDBS to standard of care cDBS that had been optimized by a movement disorders neurologist over several months. This study is the first to systematically assess real-life adaptive stimulation for PD in a naturalistic context.

The additional improvement in PD motor signs using aDBS was achieved by delivering more total electrical energy compared to cDBS in all patients. These systems therefore delivered greater stimulation during hypokinetic periods (e.g., with bradykinesia) than would be tolerated during cDBS. These results differ from most laboratory-based studies of aDBS whose goal has been reducing total electrical energy^24, 26^. The recent commercialization of rechargeable pulse generator batteries diminishes the clinical importance of conserving electrical energy.

### Limitations

The sample size was five brain hemispheres within three patients. While the group included a variety of residual motor signs and robust individualized N-of-1 statistics, the generalizability of the results may be limited. The aDBS algorithms here operated on an intermediate time scale^35^; we did not implement very fast time scale “burst trimming” algorithms that have shown benefit during in-laboratory tests^24, 36^. While this study represents the longest assessment of aDBS to date, its application was limited to a duration of one month. It is likely that with progression of PD, changes in medications or lifestyle, updates to the adaptive algorithm might be needed. While having a standardized pipeline for setting many aDBS parameters mitigates the challenge of a large parameter space, configuring personalized aDBS was nevertheless labor-intensive, required physiological expertise and may not be ready for widespread dissemination. Machine learning techniques to further automate aDBS algorithm design, and indeed to periodically update algorithms based on scores from wearables or patient reported problems, may be important for translating this technique to neurology clinics outside of specialized centers^48^.

### Summary

We demonstrate for the first time the benefit of aDBS to improve residual motor signs that persist in the setting of clinically optimized standard-of-care cDBS. aDBS improved the duration of patients’ most bothersome motor signs without aggravating other motor and non-motor symptoms, and improved patients’ quality of life. For aDBS algorithms designed to reduce residual motor fluctuations, we identified STN and cortical finely-tuned gamma oscillations, entrained at half stimulation frequency, as optimal control signals. The results were achieved by employing data–driven neural biomarker identification, controlling for independent effects of stimulation amplitude. Our findings highlight the benefit of multi-site brain recordings and at-home neural recordings with wearable monitors for configuring aDBS algorithms and have potential to inform the development of aDBS for other neuropsychiatric conditions.

## 5. Methods

### Patient evaluation and DBS device

#### Patients

We recruited three patients with PD from a population undergoing DBS implantation for motor fluctuations (male, age range: 57-68 years, disease duration: 7-12 years, pre-surgery off-medication Movement Disorder Society Unified Parkinson’s Disease Rating Scale [MDS-UPDRS]-III scores: 30-49). A movement disorders neurologist evaluated and confirmed the diagnosis of PD based on established diagnostic criteria and a neuropsychologist excluded significant cognitive impairment or untreated mood disorders. The inclusion criteria comprised motor fluctuations characterized by prominent rigidity and bradykinesia in the off-medication state, baseline off-medication MDS-UPDRS-III scores between 20 and 80, a greater than 30% improvement in MDS-UPDRS-III scores with medication compared to the off-medication state, and the absence of significant cognitive impairment (Montreal Cognitive Assessment score of 20 or above). Patients provided written consent in accordance with the Declaration of Helsinki. The Institutional Review Board of the University of California San Francisco gave ethical approval for this work. The study was registered on ClinicalTrials.gov (NCT03582891). The study protocol and the IDE application (G180097) are available through the Open Mind initiative (https://openmind-consortium.github.io).

#### Surgical procedure and DBS device

All patients underwent bilateral placement of cylindrical quadripolar deep brain stimulator leads (Medtronic model 3389) into the STN and bilateral quadripolar paddles (Medtronic model 0913025) into the subdural space over the sensorimotor cortex (Fig. 1a-b, Extended Data Fig. 1). The leads were connected to an investigational sensing-enabled implantable pulse generator (Medtronic Summit RC+S model B35300R) that was placed in a pocket over the pectoralis muscle bilaterally so that each pulse generator was connected only to ipsilateral leads. STN leads were initialized as contacts 0 to 3 (0 was the deepest contact), and cortical leads were initialized as contacts 8 to 11 (8 was the most posterior contact). Two months after surgery, the locations of the leads were verified using postoperative computed tomography (CT) scans. A more detailed account of the surgical implantation can be found in a previous publication^9^.

Summit RC+S is an investigational rechargeable bidirectional neural interface. It is capable of streaming four bipolar time domain channels simultaneously while providing standard therapeutic stimulation on up to two quadripolar leads. It can also perform aDBS using fully-embedded algorithms. The applications for sensing and aDBS were written in our laboratory in the device’s application programming interface, comply with FDA regulations for medical device software (CFR 820.30) and are available at https://openmind-consortium.github.io. Each RC+S system employs radiofrequency telemetry to establish wireless communication with an external compact relay device, which in turn transmits data to a Windows-based tablet using Bluetooth technology within a range of up to 12 m. This setup facilitates the capture of local field potentials and electrocorticography from a maximum of four bipolar electrode pairs, enabling continuous sensing during stimulation for up to 30 hours before requiring a recharge. The tablet is situated at patients’ homes and hosts custom software that allows for remote adjustment of streaming parameters and embedded adaptive DBS algorithms via an interface only accessible to researchers. Patients are able to initiate and stop streaming and report both medication intake and motor symptoms using a separate patient user interface. Further details on device characteristics are outlined in previous publications^5, 9, 33^.

#### Lead Reconstruction

Electrode positions were reconstructed by linearly coregistering postoperative CT images to preoperative T1-weighted 3T magnetic resonance imaging (MRI) scans through rigid Euclidean transformation (Extended Data Fig. 1 a-d). The LeGUI toolbox (Version 1.2)^49^ was used to perform automated correction for brain shifts^50^ and to localize electrodes to the MRI-rendered pial surfaces (Extended Data Fig. 1 e-g). Depth lead positions were reconstructed using the Lead-DBS toolbox (Version 2.6)^51^ and, when necessary, the PACER method^52^ was used to correct for brain shifts. For group analysis, electrode locations were normalized into Montreal Neurological Institute (MNI) space and STN leads were visualized using the DISTAL atlas^53^ (Fig. 1b).

### Optimization of continuous DBS

For each patient, cDBS was optimized before initiating aDBS. Optimization was performed by an independent movement disorder neurologist over a range of 11-31 (mean±standard deviation 22±10) months, with 5-10 (7.0±2.65) clinician visits. In addition, patients were allowed to make adjustments within a range of 1.6-4.0 mA (mean±standard deviation amplitude span: 2.9±0.9) at home (i.e., self-optimize stimulation). Clinical optimization was attempted first with “sense friendly” contact configurations (monopolar stimulation at contacts 1 and/or 2) but clinicians were allowed to use non-sense friendly stimulation montages if those proved clinically superior. Further, the clinical neurologist modified medications as needed (Fig. 1c). The patient’s participation in our study therefore did not constrain the clinical optimization of cDBS, except for ensuring the DBS system was set to the same stimulation frequency on both sides (to avoid sensing artifacts generated by the presence of two systems providing stimulation at different frequencies)^54^.

### Seven-step pipeline to design and implement adaptive DBS: overview

We devised an individualized, seven step data-driven pipeline to implement aDBS to treat residual motor fluctuations persisting after clinical optimization of cDBS (Fig. 2). We first identified residual bothersome motor signs on optimized cDBS (step 1). We established appropriate high and low limits for aDBS that were typically 0.5-1.0 mA higher or lower (respectively) than the optimized cDBS amplitude (step 2). We determined neural signals or “biomarkers” that best correlated with residual motor signs (steps 3 and 4). To identify reliable biomarkers of motor state for real-world aDBS, it was critical to study neural data collected across the range of medication effects and stimulation amplitudes that would be used during aDBS, and to identify signals modulated more by the patient’s underlying motor state than by the current stimulation amplitude. This was accomplished using in-clinic (step 3) and at-home (step 4) neural recordings in which both medication state and stimulation state varied. The at-home data streaming step was important to ensure that biomarkers identified in idealized, investigator-controlled conditions in the clinic could function in naturalistic settings. We then established appropriate control parameters to adjust stimulation amplitude in response to neural signals (steps 5 and 6). Finally, patients underwent a blinded, randomized comparison between cDBS and aDBS over a month per condition, conducted during patients’ normal lives (including work and travel) on the schedule of medications established during cDBS optimization (step 7).

#### Identification of patients’ residual motor signs on clinically optimized cDBS

Patients identified their most bothersome persistent motor problem while on clinically optimized cDBS (e.g., bradykinesia) in collaboration with a movement disorder neurologist (Fig. 1c and Fig. 2). Additionally, the most bothersome symptom in the opposite medication state (e.g., dyskinesia) which limited the therapeutic window during cDBS was identified. The goal of aDBS was to improve the most bothersome symptom without exacerbating the opposite symptom. For one patient, residual bothersome motor fluctuations on optimized cDBS were restricted to the right side of the body; therefore, we developed a unilateral adaptive algorithm for the left hemisphere (pat-1). The remaining two patients received bilateral, independent, adaptive stimulation algorithms (pat-2 and pat-3).

#### Determining individualized stimulation amplitude limits

We calibrated stimulation amplitudes for each patient, both on- and off-dopaminergic medication–in their hyper- and hypokinetic states, respectively–to determine the optimal amplitude limits for symptom control to be used for aDBS (Fig. 5a). Specifically, a movement disorders neurologist defined the low stimulation amplitude as the amplitude that mitigates adverse effects in the hyperkinetic state without causing breakthrough hypokinetic symptoms. Similarly, the high stimulation amplitude was identified as the amplitude that effectively manages hypokinetic symptoms, such as bradykinesia, while avoiding DBS adverse effects, such as dysarthria.

#### Monitoring motor signs

During in-clinic recordings (step 3), motor signs were assessed by a clinician using standardized rating scales. For biomarker identification at-home (step 4) and symptom monitoring during adaptive testing (step 6, step 7), we chronically monitored symptoms using wristwatch-style wearable monitors on each wrist (Parkinson’s KinetiGraph®, PKG®, Global Kinetics). These wearables employ a proprietary algorithm^30^ to provide validated scores of bradykinesia and dyskinesia in two-minute intervals. By synchronizing the neural data offline with these scores, we established brain-behavior correlations with high temporal resolution. To validate wearable outcome measures in our cohort and determine the correspondence between wearable scores and troublesome symptoms in each individual, patients completed motor diaries every 30 minutes during at least two days while wearing the monitors. Because the completion of motor diaries is effortful and difficult to maintain during normal daily activities, we thereafter relied on wearable data when applicable for symptom analyses to reduce patient burden.

#### Neural recordings

At multiple times during steps 3-7, patients streamed neural data and reported medication intake and motor symptoms using the patient graphical user interface on the streaming tablet. We sampled neural time series data from one subcortical and two cortical leads in bipolar configuration. We used sampling rates of 250-500 Hz to minimize data loss that may occur at higher sampling rates^9^. STN LFPs were recorded in a bipolar configuration with contacts immediately adjacent to the stimulating cathode, providing common mode rejection of the stimulus artifact during active stimulation. Cortical recordings were performed in non-overlapping bipolar pairs with cortical montage 1 referring to the anterior montage which has at least one or both electrode contacts on precentral gyrus, and cortical montage 2 referring to the posterior montage that has at least one or both electrode contacts on postcentral gyrus. Data were encrypted and uploaded to a secure cloud environment.

### Biomarker identification: details of steps 3 and 4

#### Data collection procedure

In step 3, we recorded neural data off- and on-dopaminergic medication in-clinic, thus in provoked hypo- and hyperkinetic states, during stimulation at the previously identified low and high stimulation amplitudes. We performed off-medication recordings after at least 12 hours of overnight medication withdrawal. For each of the four combinations of medication state and stimulation amplitude, we obtained 26.5±5.84 (mean±standard deviation) minutes of recordings while patients performed a standardized set of activities including walking, resting, speaking, and eating and the MDS-UPDRS-III. We then confirmed the applicability of the in-clinic biomarker in a real-life setting at patients’ homes (step 4). To that end, patients recorded neural data during their regular daily activities while we monitored symptoms using wearable devices and motor diaries. Patients recorded at least two levodopa medication cycles of neural data at both low and high stimulation amplitudes. During streaming days, we recorded neural time-series data and onboard power of the device in the frequency bands of interest defined based on in-clinic recordings.

#### Neural data analysis

We performed all analyses using MATLAB 2021a (The Mathworks, Natick, MA, USA) and the FieldTrip toolbox^55^. We first computed the power spectral density of non-interrupted time segments of the neural signal using Welch’s method with 1 s windows and 95% overlap mimicking RC+S embedded system capabilities. We calculated power spectral density over 2-100 Hz in non-overlapping 2.5 s epochs of time domain signals using a 1 Hz spectral resolution. The RC+S device streams data to a laptop computer and time stamps the neural data using the computer clock, which we used to synchronize neural data with wearable monitors.

#### Statistical analysis for identification of biomarkers: overview

We employed a data-driven approach integrating non-parametric statistics and machine learning to identify patient-specific neural biomarkers (Extended Data Fig. 4). By analyzing the entire available frequency range in both the STN and sensorimotor cortex (2-100 Hz), we identified physiological signals that reliably predicted each patient’s most bothersome or opposite motor symptom. We ensured these physiological signals were not independently influenced by stimulation changes (Extended Data Fig. 5) in a direction that could lead to undesired cyclic behavior in the control system unrelated to symptom fluctuations^33^. For the analysis of in-clinic data, we assessed the main effects of medication and stimulation, as well as their interactions, using a 2×2 factorial design. To this end, we employed a non-parametric cluster-based permutation analysis—a widely-used neuroscientific method that operates free of a priori assumptions regarding the data distribution. It further allows exploration of the complete frequency spectrum while effectively controlling for multiple comparisons^56^. At-home recordings provided a rich dataset with long-term neural time series and continuous symptom monitoring in patients’ naturalistic environment, therefore we did not have to rely on decoding medication states. We instead used stepwise-linear regression to predict bothersome/opposite symptoms for patients with continuous symptom monitoring (i.e,. upper limb symptoms measurable by wrist-watch style wearables), and a linear discriminant analysis (LDA) based method for binary-classified (rather than continuously scaled) symptoms, i.e., presence or absence of lower limb dystonia. We also expanded this LDA method to all patients by identifying data-driven mappings between continuous wearable scores and self-reported symptom labels. Both the non-parametric cluster-based analysis and the linear stepwise regression explicitly modeled the contributions of stimulation effects, whereas this effect was implicitly addressed with the LDA by using equal distribution of stimulation amplitudes within the training data sets.

#### Statistical analysis of in-clinic data: details

To implement non-parametric cluster-based permutation, we ranked the test statistic (here sum of *t*-values) of the empirical in-clinic data within a permutation distribution obtained by randomly assigning condition labels to each data segment. We matched the amount of data for each condition (medication and stimulation state) by drawing equally as many samples from each of the four conditions (1000 random draws), and thereafter ranked clusters among 1000 surrogates and assessed the main effects of medication (across stimulation conditions) and stimulation (across medication states), as well as their interaction on power, using two-sided statistical tests^57^. We determined effect size using Cohen’s *d* and selected the neural signal with the largest main effect of medication as the optimal control signal. We ensured the neural signal for adaptive control was unaffected by independent contributions of stimulation to prevent adverse impact on the performance of the adaptive algorithm (Extended Data Fig. 5). This involved confirming the stimulation amplitudes for adaptive control did not produce any significant amplitude augmentation of on-state biomarkers (neural signals that increase after medication intake or during hyperkinetic states), nor any significant amplitude diminution of off-state biomarkers (neural signals that decrease after medication intake and increase during hypokinetic states). Statistically this is expressed as positive or negative effects of stimulation amplitude, respectively. We assessed effects in the STN and the two cortical regions separately and Bonferroni-corrected *p*-values for multiple comparisons. We used an alpha level of 0.05.

#### Statistical analysis of at-home data: details

For at-home data, we used two analysis approaches. For patients 1 and 3, continuous wearable data tracked their most bothersome/opposite symptoms. Thus, we calculated a linear stepwise regression using data from all three brain regions and the frequency spectrum from 2-100 Hz as predictors for the most bothersome symptom (pat-1: bradykinesia), or the opposite symptom that limits the therapeutic window (pat-3: dyskinesia). We *z*-scored predictors and modeled stimulation amplitude as an additional feature in order to extract independent contributions of power bands to symptom prediction. We excluded episodes in which the wearable score indicated an immobility level predictive of sleep (bradykinesia score >80^58^) or the monitor was labeled as being off-wrist. However, as we could not rule out neural data being recorded during daytime naps completely, we excluded very low frequencies as predictors that can be confounded by sleep (2-4 Hz)^59^. Using this method, we identified the strongest predictor of patients’ symptom states. We Bonferroni-corrected *p*-values for multiple comparisons (289 predictors) and used an alpha level of 0.05.

For patient 2, their most bothersome symptom, i.e., lower limb dystonia, could not be measured with wearables. Instead, this patient completed motor diaries at least every 30 minutes indicating the presence or absence of lower limb dystonia during neural streaming. We used an LDA based method to identify patient-specific neural signal biomarkers that maximized discriminability between the presence and absence of the most bothersome symptom. Initially, we identified the top five candidate 1 Hz power bands from the neural signal spectra. These bands were selected based on their ability to maximize the area under the receiver operating curve (AUC) through a 1000-repetition Monte Carlo cross-validation. The validation process involved randomly drawing an equal number of data points at both stimulation amplitudes, with a 10% subset used for testing. Within each fold of the Monte Carlo cross validation, the optimal band widths for these potential biomarkers were optimized as a hyperparameter via a nested 10-fold cross validation. The final power band width for each of the five potential biomarkers was chosen as the aggregation of 1 Hz bands that were present in 99% of the Monte Carlo folds.

Given the RC+S device’s embedded aDBS capabilities rely on discrete classification of neural signal biomarkers, we additionally assessed patient 1 and 3’s at-home data using the LDA method. To implement this for these patients, we used a nonlinear optimization (MATLAB *fminbnd* function) to identify patient-specific dichotomizing boundaries that best mapped the continuous wearable symptom scores into binary symptom labels (i.e., symptom present versus not). Data used for this optimization drew from days when patients provided simultaneous motor diaries and wearable symptom metrics. The optimization value function was the F1 score for predicting the motor diary symptom label from the contemporaneous wearable score’s relationship to the dichotomizing boundary (whether above or below). Boundary values calculated for dyskinesia scores ranged from 8.5-14.1 and bradykinesia scores ranged from 11.7-21.7.

#### Offline assessment of biomarker performance

Given the preponderance of literature highlighting STN beta power’s potential as a neural signal biomarker for aDBS, we compared the offline prediction of medication state/symptoms (Step 3: Cohen’s *d*, Step 4: regression statistics and LDA AUC) by our identified biomarkers to that of subthalamic beta power bands. Beta bands used for this comparison were identified by repeating the above analyses for Steps 3 and 4 while constraining the search to subthalamic frequency bands within the beta range (13-30 Hz).

Because the RC+S system is capable of using up to four neural biomarkers as inputs into its aDBS systems, we also assessed the potential additional benefit of using beta band biomarkers in conjunction with our biomarkers. LDA identified the optimal linear combination of the two subject-specific biomarkers (data-driven and STN beta-constrained) to predict the presence/absence of bothersome symptoms. The significance of AUC changes resulting from using STN beta power (identified from Step 3) in addition to the data-driven frequency band was assessed by comparing the resulting AUC to a surrogate distribution of AUC values. The surrogate distribution was produced by LDA classifiers using the data driven band and one of 1000 randomly selected power bands of analogous width to the beta band, but unconstrained to anatomic location or frequency range.

### Optimization of adaptive parameters: details of steps 5 and 6

The stimulation contact, frequency, and pulse width remained consistent between aDBS and cDBS, with only the stimulation amplitude varying in response to estimates of the patient’s clinical state (i.e., presence of symptoms). During steps 5 and 6, we identified and refined additional parameters that governed both how clinical state estimates were predicted from data-driven biomarkers and the system’s temporal dynamics. To briefly summarize these parameters: the embedded aDBS platform uses windows of time domain data with frame shifts at specified intervals to compute the fast Fourier transform (FFT). Thereafter, it calculates the input signal for the control algorithm, the linear detector (LD), by averaging the signal across a researcher-specified number of FFT windows, known as the update rate. The temporal resolution of the LD is therefore determined by the sampling frequency, FFT window, FFT interval, and update rate. At each update, the input signal is compared to researcher-identified thresholds in order to predict the patient’s clinical state (i.e., whether they are having a symptom that requires stimulation adjustment). The device allows up to two thresholds to be set per detector, corresponding to three states (Extended Data Fig. 6c-e). Changes in stimulation are then governed by a look-up table for each clinical state. The temporal dynamics of the final adaptive algorithm can also be influenced by the tolerated stimulation ramping time (e.g., 0.1 mA/s ramp rate for 2.0 mA increase in stimulation amplitude=20 seconds) and the onset and termination rate, which define the number of “updates” the LD is required to be above or below the threshold to result in a change of stimulation amplitude.

In step 5, we performed supervised testing of an initial adaptive algorithm to find individualized ramp rates and assure patient comfort with stimulation amplitudes (e.g., avoiding paresthesia). We used the predictive power bands identified in the previous steps and chose thresholds to trigger state changes based on visual inspection of the control signal. In this step of our workflow, we were interested in many state transitions during the brief testing period in-clinic in order to test adaptive parameters during stimulation changes for aDBS. We thus used an aDBS algorithm with relatively fast temporal dynamics that differed from algorithms implemented at home (update rate=1.5 s, onset and termination period=0). After identifying optimal ramp rates during which patients did not perceive stimulation changes, we moved on to unsupervised at-home testing of adaptive algorithms in step 6, the aDBS optimization phase.

During step 6, patients first streamed the selected neural biomarker as an on-board power band with the fastest temporal dynamics possible for several days on cDBS alongside symptom monitoring to obtain an initial set of aDBS parameters. We monitored symptoms using wearables and patient comments on symptom onset and offset in the patient-facing app on the tablet. Thereafter, we conducted brief 24-hour unblinded tests of aDBS algorithms, in which we refined thresholds, onset and termination periods and, if necessary, stimulation amplitudes based on aDBS’ effects on symptoms (see *Mitigating noise and artifact effects*). Subsequently, we performed 24-hour blinded testing and assessed aDBS performance on daytime symptoms and sleep quality using a daily symptom application as an outcome measure (see *Procedure and outcomes measures*). We assured that aDBS algorithms did not decrease sleep quality, in which case we would consider developing a sleep-aware algorithm by adding a second neural biomarker for sleep, e.g., cortical low delta power^11^. A detailed summary of the final adaptive stimulation parameters can be found in Extended Data Fig. 6b. A comprehensive set of starting parameters for algorithms on a similar time scale of minutes to hours is summarized in Extended Data Fig. 6a.

#### Thresholds for state transitions

We identified LD thresholds separating times with and without the most bothersome motor sign in step 6. Preliminary thresholds were calculated based on several days of streamed LDs during cDBS and thereafter fine-tuned based on results from adaptive testing. We used a binary classification algorithm aimed at balancing sensitivity and specificity while considering patient preference. We used a receiver operating characteristic curve (ROC) to identify the threshold points on the ROC curve for optimal trade-off between true and false positive rate^60^. Initially, we used a single threshold algorithm for all patients and continuously monitored neural data and symptoms (Extended Data Fig. 6c).

#### Mitigating noise and artifact effects

To mitigate noise that could lead to erroneous stimulation changes, we employed three strategies. Firstly, we reduced inherent noise by decreasing temporal resolution, i.e., smoothing the LD by increasing the update rate. To ensure adequate responsiveness of the algorithm, we enforced a maximal averaging of one minute. If the biomarker signal-to-noise ratio still resulted in erroneous threshold crossings, we introduced a middle state as a noise buffer zone, in which stimulation amplitude remained constant (Fig. 5c, Extended Data Fig. 6e). Alternatively, if we observed brief and rare artifacts, we increased the onset and termination duration. Each of these parameter changes was guided by the patients’ symptoms and satisfaction as assessed by questionnaires and wearable devices for several days. To mitigate the potential effects of artifact produced by stimulation ramping^33^, a detector blanking period exceeding the update rate by one second was implemented such that the algorithm ignored neural signal spectral content calculated during a change in stimulation. To mitigate the potential for electrocardiogram artifacts seen in other sensing DBS devices^61, 62^, charge at the tissue-electrode interface was actively redistributed after the stimulation impulse was delivered (“active recharge”).

### Blinded randomized comparisons of continuous and adaptive DBS

#### Procedure and outcome measures

In our final step 7, we conducted a blinded, randomized comparison between the effects of aDBS and clinically optimized cDBS on motor signs. Both stimulation conditions were applied for blocks of 2-4 days over the course of one month per condition at patients’ homes. To assess outcomes, we utilized patients’ daily symptom diaries and wearable data. We asked patients to complete the daily symptom diary implemented as a custom electronic questionnaire every night before bedtime. The questionnaire focused on the total number of hours spent with symptoms, symptom severity, and a quality of life (QoL) score validated for daily assessment of health-related QoL (EQ-5D)^63^. Evaluated symptoms included the most bothersome and opposite symptom as well as a range of additional common motor symptoms (bradykinesia, dyskinesia, tremor, dystonia, dysarthria, and gait disturbance; Extended Data Fig. 8). We included one question asking patients to rate their quality of sleep. Symptom severity and sleep quality were rated on a scale from 1-10. We also assessed nonmotor symptoms (depression, anxiety, apathy, and impulsivity) using the wording of the four-point scale modeled off the MDS-UPDRS-I^64^. In addition, we inquired daily about patients’ perceived stimulation condition (aDBS/cDBS) and the underlying rationale. They were given the choice to attribute their experience to unusual sensations related to changes in stimulation, as well as improvements or worsening of motor symptoms. One patient perceived paresthesias during the beginning of the testing period. We therefore excluded these data points and reduced the aDBS ramp rate for future testing.

Evaluation of the wearable monitor scores focused on quantifying changes in motor fluctuations. Logs from the DBS device during both aDBS and cDBS days indicated the time stamps at which the LD changed its estimate of clinical state, which allowed us to label wearable scores as being present during hypokinetic or hyperkinetic periods. Within each day, bradykinesia and dyskinesia scores were averaged for each clinical state. Our primary metric for assessing effect on motor fluctuations between cDBS vs aDBS (fluctuation score) was the within-day difference in symptom (both bradykinesia and dyskinesia) magnitude between hypokinetic and hyperkinetic states. We also assessed the difference in bradykinesia score magnitude during hypokinetic states between cDBS and aDBS days, and the difference in dyskinesia score during hyperkinetic states. Each brain hemisphere/contralateral hemibody pair were assessed independently.

Furthermore, using logs from the DBS device, we calculated the percentage of each aDBS day and night spent at the high and low stimulation amplitudes and the daily average consecutive duration of each stimulation amplitude in hours. Finally, we calculated the total electrical energy delivered (TEED) per second during both stimulation conditions assuming an impedance of 1000 Ω for both day and night time^65^.

#### Statistics of adaptive versus continuous DBS effects

We performed within-subject statistics to compare the clinical effects of aDBS to cDBS. Given smaller sample sizes, we used nonparametric statistical tests. For each subject, we used the Wilcoxon rank sum test to assess planned comparisons of the nightly questionnaire data for the percentage of awake hours with the patient’s bothersome symptom, the percentage of awake hours with the patient’s opposite symptom, and quality of life scores. Similar methods were used to evaluate within subject/hemisphere differences in wearable monitor motor fluctuation metrics for the bothersome and opposite symptom (in the two patients with upper limb symptoms). For questionnaire data in addition to these planned comparisons (such as other motor symptoms and non-motor symptoms) and other wearable metrics, we used the Wilcoxon rank sum test adjusting for multiple comparisons using the false discovery rate procedure. For patient 3, who had bilateral aDBS systems, the wearable metrics for the two hemibodies were corrected separately for multiple comparisons.

## Data Availability

All data produced in the present study are available upon reasonable request to the authors

## 7. Acknowledgements

The study was supported by NINDS UH3NS100544. CRO, SC, LHH, and JY were funded by the Parkinson Fellowship of the Thiemann Foundation, NINDS F32NS129627, NINDS R25NS070680, and the TUYF Charitable Trust Fund respectively. Research reported in this publication was supported by the National Institute Of Neurological Disorders And Stroke of the National Institutes of Health under Award Number K23NS120037 & R01 NS131405-01. We acknowledge the OpenMind Consortium for technical support throughout the project (funded by NINDS U24NS113627). We thank T. Wozny for lead localization, C. Smyth for technical contributions, and Ken Probst for medical art (Fig. 1a). The content is solely the responsibility of the authors and does not necessarily represent the official views of the National Institutes of Health.

## Notes

### Competing Interest Statement

SL has received honorarium from Medtronic and is a paid consultant for Iota Biosciences.

### Clinical Trial

NCT03582891

### Clinical Protocols

https://osf.io/cmndq

### Author Declarations

The Institutional Review Board of the University of California San Francisco gave ethical approval for this work.

