## Extended\Supplemental Materials for "Personalized chronic adaptive deep brain stimulation outperforms conventional stimulation in Parkinson’s disease"

### Extended Data Figures

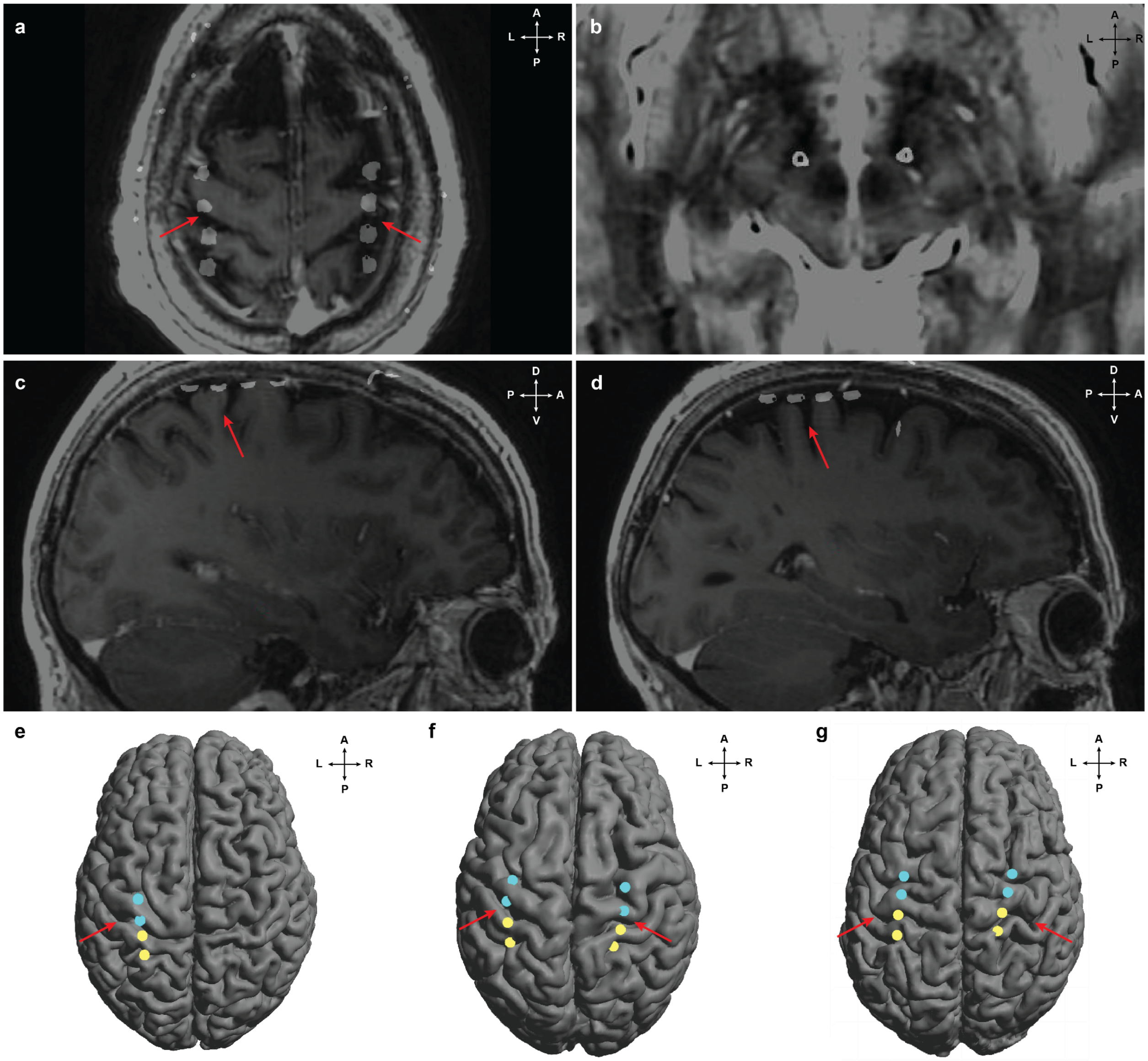


**Extended Data Fig. 1. Localization of leads over sensorimotor cortex and within subthalamic nucleus in native space. a-d,** Example localization of cortical and subcortical leads in patient 2, generated by fusing postoperative CT with preoperative MRI scans. Contacts appear as white CT artifacts due to metal content. **a**, Cortical leads on axial T1-weighted MRI through the vertex. **b**, STN leads on axial T2-weighted MRI through the region of the dorsal STN, 3 mm inferior to the intercommissural plane. **c-d**, Cortical leads on oblique sagittal T1-weighted MRI passing through the long axis of the lead array in left **(c)** and right **(d)** hemispheres, respectively. **e-g,** Location of cortical leads overlayed on 3D reconstruction of cortex rendered using LeGUI. Electrodes used in cortical montages 1 and 2 are shown in cyan and yellow, respectively. For patient 1 **(e)**, and 2 **(f)**, montage 1 and 2 covered the pre- and postcentral gyrus, respectively. For patient 3, montage 1 included one electrode on the middle frontal and one on the precentral gyrus. Montage 2 comprised one pre- and one postcentral electrode. In all figures, red arrows indicate the location of the central sulcus.

**a**

| | hemi-sphere | main effect medication<br>brain region | <br>frequency band (Hz) | <br>Cohen's d | <br>p-value | main effect stimulation<br>frequency band (Hz) | <br>Cohen's d | <br>p-value | interaction effect<br>frequency band (Hz) | <br>Cohen's d | <br>p-value |
|---|---|---|---|---|---|---|---|---|---|---|---|
| Patient 1 | L | cortical montage 1 | 63-67 | 2.56 | p<0.001 | 59-63<br>67-70 | 0.83<br>0.7 | p<0.01<br>p<0.01 | X | X | X |
| Patient 2 | R | STN | 64-66 | 1.14 | p<0.001 | X | X | X | 64-66 | -1.25 | p<0.001 |
| Patient 2 | L | STN | 64-66 | 0.87 | p<0.001 | 65 | -0.61 | p<0.001 | 64-66 | -0.84 | p<0.001 |
| Patient 3 | R | cortical montage 2 | 64-66 | 0.54 | 0.042 | X | X | X | X | X | X |
| Patient 3 | L | no significant effects found | | | | X | X | X | X | X | X |

**b**

| | hemi-sphere | main effect medication<br>brain region | <br>frequency band (Hz) | <br>Cohen's d | <br>p-value |
|---|---|---|---|---|---|
| Patient 1 | L | STN | 20 | -0.32 | 1 |
| Patient 2 | R | STN | 10-16 | -0.98 | p<0.01 |
| Patient 2 | L | STN | 17-18 | -0.3 | 0.08 |
| Patient 3 | R | STN | 26 | -0.18 | 1 |
| Patient 3 | L | STN | 14-18 | -1.32 | 0.64 |

**c**

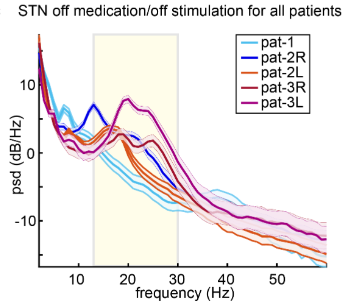


**d**

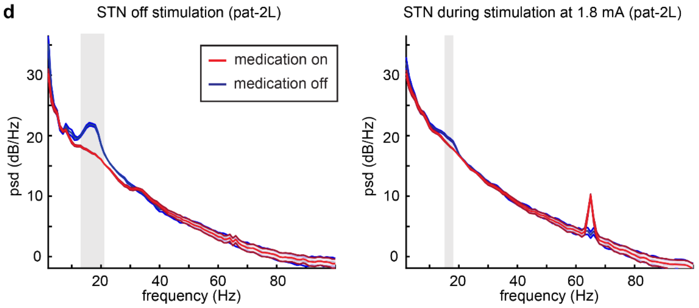


**Extended Data Fig. 2. Neural biomarkers of medication effects identified in-clinic.** All tables show the results from our within-patient non-parametric cluster-based permutation analyses using in-clinic recordings during two medication states (off vs. on) and stimulation conditions (low vs. high stimulation amplitude). *P*-values were Bonferroni-corrected for multiple comparisons. **a,** Statistics for the largest main effect of medication, stimulation, and their interaction for each patient and hemisphere when searching the whole frequency space (2-100 Hz) across brain regions. Frequencies represent the center frequency of 1-Hz wide power spectral density bins. For all three patients (four hemispheres), we found that finely-tuned gamma power in the STN or cortex was the best predictor of medication state. Positive Cohen's *d* values highlight that the neural biomarker was higher during on-medication states. For patients 2 and 3, we did not find overlapping potentially problematic positive stimulation effects on biomarkers identified for a main effect of medication (for an illustration of problematic stimulation effects on adaptive algorithms see Extended Data Fig. 5). For patient 1, we excluded 63 and 67 Hz from the subsequently used control signal due to intersecting positive stimulation effects. **b,** When constraining the anatomic location and frequency space to STN beta oscillations (13-30 Hz), STN beta power was only predictive for medication state in one hemisphere and smaller in effect size than cortical/STN gamma oscillations for all patients. **c,** Power spectral density in the STN based on in-clinic recordings off medication and off stimulation for all five hemispheres. All but one hemisphere (pat-1) exhibited a peak in the beta frequency band (illustrated in yellow). **d,** Example of the suppressive effect of DBS on STN beta oscillations leading to beta power being a less adequate biomarker during active stimulation (pat-2L, all data collected during the same in-clinic recording session). Off stimulation, the spectral peak in the beta frequency range was suppressed by medication (13-21 Hz, Cohens' $d$=-1.09, $p$<0.001). However, this medication effect diminished during active stimulation, even at low stimulation amplitudes (1.8mA, largest effect in the beta band: 15-18 Hz, Cohens' *d*=0.31, *p*=0.026). Data are corrected for stimulation-induced broadband shifts.

**a**

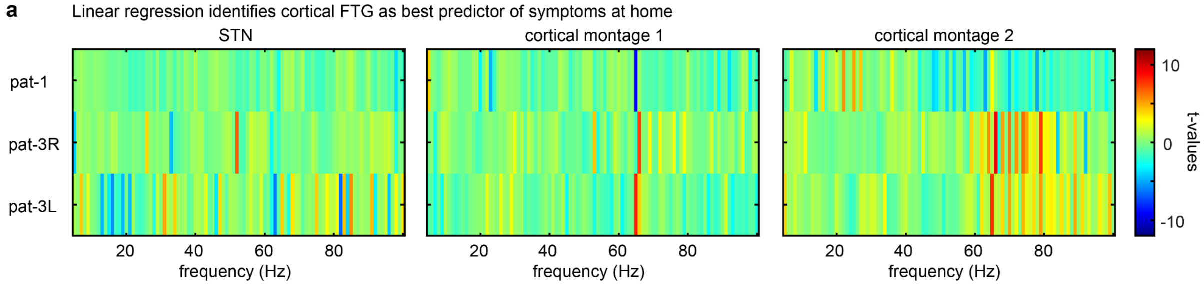


**b**

| | symptom | Linear regression: brain region | Linear regression: frequency band (Hz) | Linear regression: beta value | Linear regression: t-value | Linear regression: p-value | LDA: brain region | LDA: frequency band (Hz) | LDA: AUC |
|---|---|---|---|---|---|---|---|---|---|
| pat-1 | bradykinesia | cortical montage 1 | 65 | -2.89 | -9.12 | p<0.0001 | precentral gyrus | 65 | 0.69 |
| pat-2R | lower limb dystonia | no continuous symptom monitoring available | | | | | STN | 66-67 | 0.71 |
| pat-2L | lower limb dystonia | no continuous symptom monitoring available | | | | | STN | 65 | 0.79 |
| pat-3R | dyskinesia | cortical montage 2 | 66-68 | 0.86 | 9.34 | p<0.0001 | pre-postcentral | 65-71 | 0.85 |
| pat-3L | dyskinesia | cortical montage 2 | 65-66 | 0.9 | 7.71 | p<0.0001 | pre-postcentral | 65 | 0.70 |

**c**

| | symptom | Linear regression: brain region | Linear regression: frequency band (Hz) | Linear regression: beta value | Linear regression: t-value | Linear regression: p-value | LDA: frequency band (Hz) | LDA: AUC |
|---|---|---|---|---|---|---|---|---|
| pat-1 | bradykinesia | STN | 21 | -0.15 | -1.53 | 1 | 28 | 0.52 |
| pat-2R | lower limb | no continuous symptom monitoring available | | | | | 13-18 | 0.65 |
| pat-2L | lower limb | no continuous symptom monitoring available | | | | | 16-22 | 0.64 |
| pat-3R | dyskinesia | STN | 26 | 0.29 | 3.77 | 0.048 | 18 | 0.54 |
| pat-3L | dyskinesia | STN | 16 | -6.27 | -5.77 | p<0.0001 | 16-18 | 0.68 |

**d**

| | symptom | Linear regression: brain region | Linear regression: frequency band (Hz) | Linear regression: beta value | Linear regression: t-value | Linear regression: p-value | LDA: frequency band (Hz) | LDA: AUC |
|---|---|---|---|---|---|---|---|---|
| pat-1 | bradykinesia | STN | 21 | -0.15 | -1.53 | 1 | 28 | 0.52 |
| pat-2R | bradykinesia | STN | 15 | -0.51 | -3.68 | 0.07 | 13 | 0.56 |
| pat-2L | bradykinesia | STN | 14 | -0.34 | -2.84 | 1 | 28-30 | 0.53 |
| pat-3R | bradykinesia | STN | 23 | -0.23 | -1.27 | 1 | 25 | 0.53 |
| pat-3L | bradykinesia | STN | 19 | 2.45 | 10.18 | p<0.0001 | 16 | 0.53 |

**Extended Data Fig. 3. Neural biomarkers of symptoms identified at-home. a**, Heatmaps of *t*-values derived from stepwise linear regressions using 1 Hz power bands between 2-100 Hz in the STN (left), cortical montage 1 (middle) and cortical montage 2 (right) to predict the bothersome/opposite symptom measured with continuous wearable monitors for patients 1 and 3. **b**, Linear regression (left) and linear discriminant analysis (LDA; right). Both methods provide converging evidence that finely-tuned gamma (FTG) power centered at half the stimulation frequency (65 Hz) in the STN and cortex optimally distinguishes hypo- and hyperkinetic symptoms. **c,** When constraining the anatomic location and frequency space to STN beta oscillations (13-30 Hz), frequency bands identified as most predictive of bothersome/opposite symptoms were less discriminative than cortical/STN gamma oscillations (all AUC<0.7). Corresponding linear regression models also resulted in smaller magnitude coefficients with only one hemisphere, which demonstrated a significant negative association with hyperkinetic symptoms (pat-3L). All *p*-values were Bonferroni-corrected for multiple comparisons (289 predictors). **d,** STN beta frequency bands were also poorly predictive of wearable bradykinesia scores (AUC<0.6), again with only one hemisphere demonstrating a significant effect in the regression model (corresponding to positive relationship with hypokinetic symptoms; pat-3L).

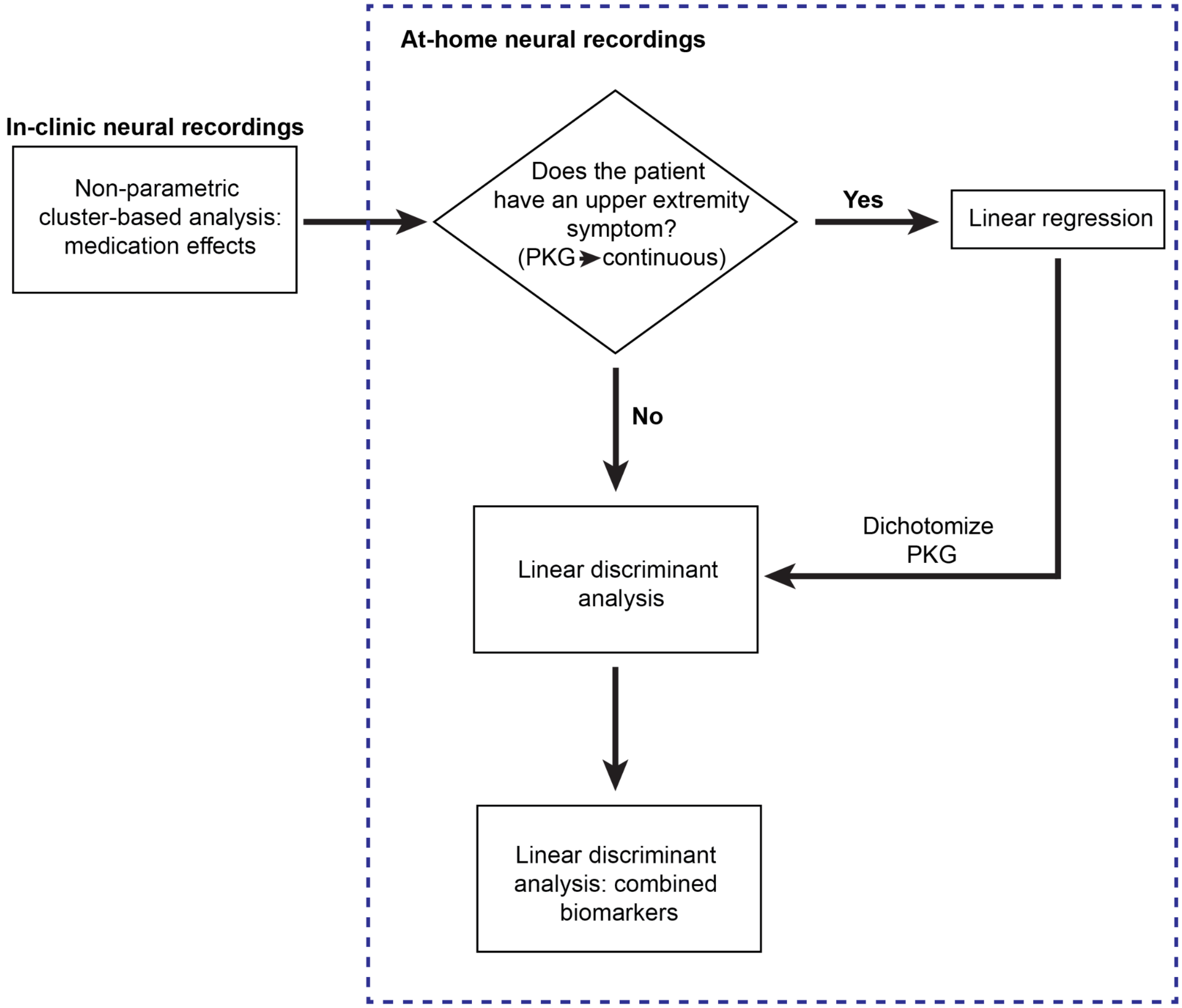


**Extended Data Fig. 4. Flowchart of biomarker identification analyses.** We identified neural biomarkers using standardized in-clinic and at-home recordings in patients' naturalistic environments. Non-parametric cluster-based permutation analysis identified candidate spectral biomarkers from in-clinic data by assessing main effects of medication state, stimulation amplitude, and the interaction. Next, the predictability of neural biomarkers as robust aDBS control signals of symptom state was tested using at-home recordings. For patients where the most bothersome symptom was monitored by a wearable device (e.g., upper extremity bradykinesia or dyskinesia), linear stepwise regression was used to take advantage of the continuous nature of the symptom measurements. The most predictive frequency bands and recording sites were selected based on *t*-values. If the patient's most bothersome symptom could not be captured by wearable monitors, the patient's motor diaries and streaming app entries instead labeled the presence of symptoms. A linear discriminant analysis (LDA) based method identified the most predictive frequency band and recording site from these discretely labeled neural signal data, as measured by the area under the receiver operating curve (AUC). We also applied the LDA-based approach to symptoms measured by wearable monitors by mapping the continuous wearable scores to discrete symptom labels using a patient-specific dichotomization. This dichotomization allowed for subsequently assessing prediction accuracy based on multiple neural biomarkers.

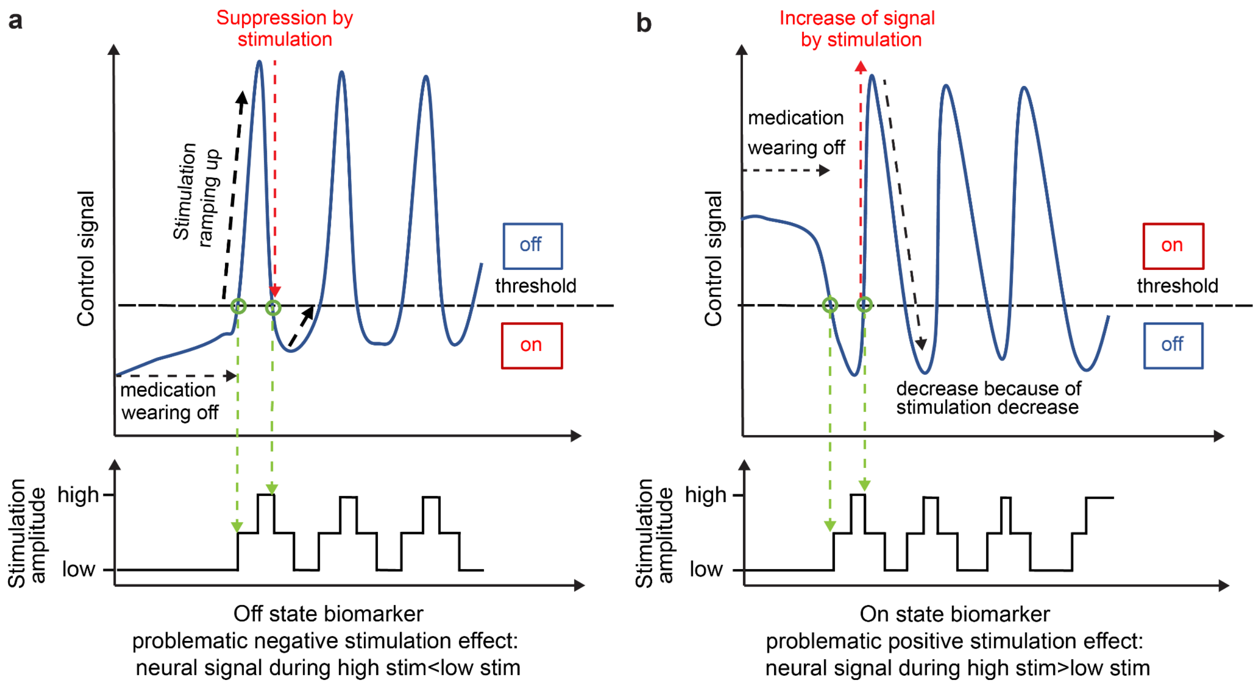


**Extended Data Fig. 5. Problematic stimulation effects on neural signals.** Two examples of self-triggering of the control algorithm caused by stimulation effects. Upper sub-panels illustrate the neural signal in blue and thresholds as dashed black lines, and the lower subpanel highlights the corresponding stimulation amplitude. **a**, Off-state biomarkers are defined as neural signals that are higher when patients experience hypokinetic symptoms (e.g., beta oscillations). The control algorithm therefore increases stimulation amplitude when the neural biomarker is high. Self-triggering may occur when increases in stimulation amplitude suppress the control signal to a greater degree than enhancements of the control signal resulting from natural medication wear-off (in our nonparametric cluster-based analysis: a negative stimulation effect). As medication wears off and the neural signal increases over a set threshold (left green circle), the resulting increase in stimulation can subsequently suppress the biomarker until it then falls below the threshold (right green circle), leading to cyclic behavior. **b**, For on-state biomarkers, as in our study, the opposite stimulation effect would be problematic. Here, control signals are higher after medication intake when patients are more prone to hyperkinetic symptoms. In aDBS algorithms like ours, stimulation amplitude increases when medication naturally wears off and hypokinetic symptoms manifest (left green circle). Self-triggering may occur when this change in stimulation amplitude leads to an artificial enhancement of the neural signal that exceeds the natural signal decrease observed during hypokinetic times, as this leads to an inappropriate passing of the threshold (right green circle). Similar to the off-state biomarker, the result is cyclic behavior of the control system, independent of patients' symptomatic state. In our nonparametric cluster-based analysis this is reported as a positive stimulation effect. For our identified on-state biomarkers in the gamma frequency range, we therefore identify spectral biomarkers without positive stimulation effects (Extended Data Fig. 2).

**a**

| | Recommeded starting control policy parameters (beginning of step 6) | | | | | | | | | |
|---|---|---|---|---|---|---|---|---|---|---|
| | control policy | lower stimulation amplitude (mA)* | upper stimulation amplitude (mA)* | threshold* | FFT interval (ms) | update rate (s) | blanking period (s)* | onset duration (# LDs) | termination duration (# LDs) | ramp rate (mA/s)* |
| data source | | result from step 2 | result from step 2 | result from step 6 cDBS streaming | | | ramping time + 1 second | | | result from step 5 |
| example | single | 1 | 4 | 200 | 50 | 10 | 4 | 0 | 0 | 1 |

*parameters based on results from previous steps

**b**

| | Control signal | | | Control policy parameters | | | | | | | | |
|---|---|---|---|---|---|---|---|---|---|---|---|---|
| | hemi-sphere | brain region | frequency (Hz) | control policy | low threshold | high threshold | FFT interval (ms) | update rate (s) | blanking period (s) | onset duration (# LDs) | termination duration (# LDs) | ramp rate (mA/s) |
| Patient 1 | L | cortical montage 1 | 64-66 | buffer | 150 | 500 | 50 | 30 | 25 | 3 | 0 | 0.1 |
| Patient 2 | R | STN | 64-66 | single | 500 | NA | 50 | 15 | 40 | 2 | 2 | 0.05 |
| Patient 2 | L | STN | 64-66 | single | 350 | NA | 50 | 15 | 40 | 2 | 2 | 0.05 |
| Patient 3 | R | cortical montage 2 | 64-69 | single | 200 | NA | 50 | 30 | 1.5 | 3 | 3 | 1 |
| Patient 3 | L | cortical montage 2 | 64-69 | single | 220 | NA | 50 | 30 | 1.5 | 3 | 3 | 1 |

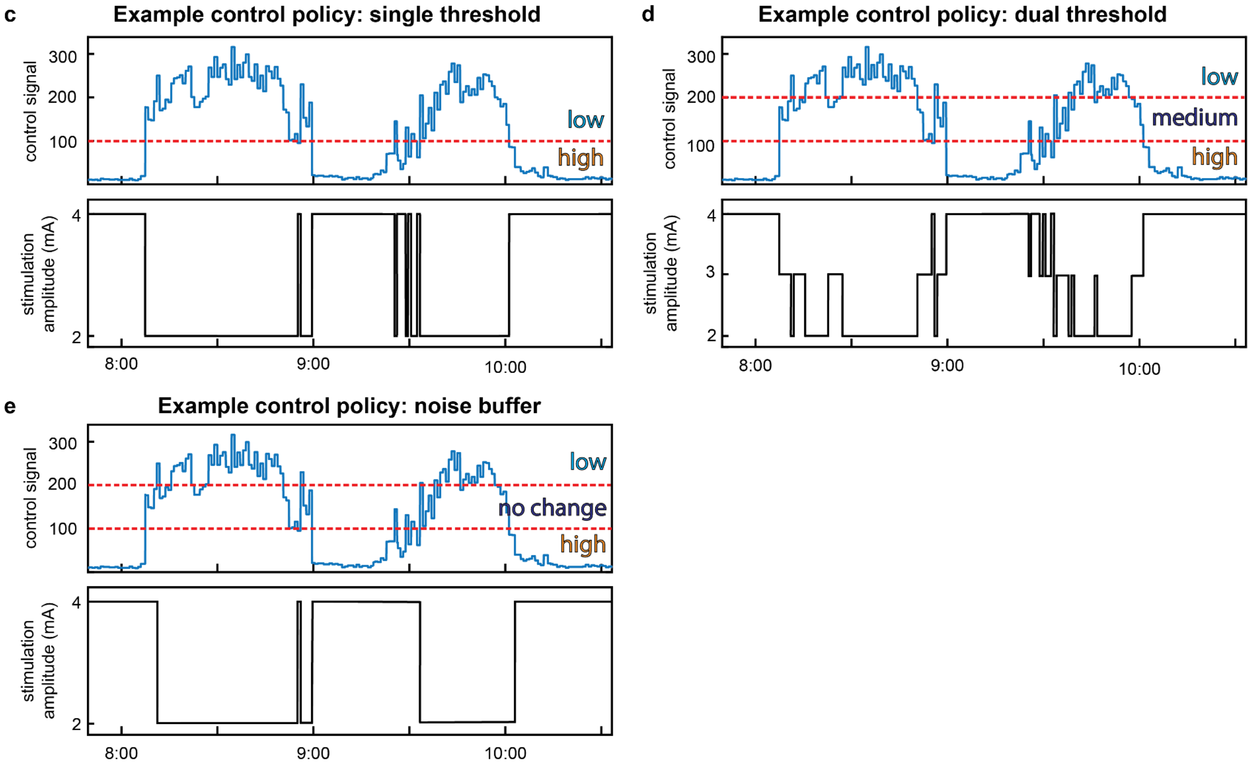


**Extended Data Fig. 6. Initial and finalized adaptive stimulation parameters and example adaptive control policies. a**, Suggested initial parameters for algorithms developed for time scales of minutes to hours, as identified during steps 5 and 6 of the pipeline. An update rate of 10 s typically provided a signal to noise ratio that allowed for a delineation between the presence and absence of the most bothersome symptom, and often improved when increasing it further. The ramp rate chosen for each patient depended on the results of step 5 (we chose an example of 1 mA/s). **b**, Detailed final adaptive stimulation parameters including control signals, thresholds, FFT interval, update rates, blanking periods, onset and termination duration, and ramp rates used for each patient and hemisphere. **c-e**, Examples of potential control policies that can be used for an adaptive algorithm using artificial data. The upper subpanels of each subfigure illustrate an on-state biomarker (blue), as used in our study, along with thresholds (red). Lower subpanels demonstrate the adjustment of stimulation amplitude based on the relationship of the neural signal to the thresholds. **c**, A single threshold control policy with two stimulation amplitudes. When the biomarker is above the threshold, stimulation amplitude decreases and once below threshold, stimulation amplitude increases. **d**, A dual threshold control policy with three stimulation amplitudes, which may be used to address three symptom states or approximate a proportional stimulation change in response to the biomarker. When the neural signal is below both thresholds, the stimulation amplitude is high (e.g., 4 mA). When the biomarker is between the two thresholds, stimulation adjusts to a middle amplitude (e.g., 3 mA). When the biomarker exceeds the second threshold, stimulation decreases to the low amplitude (e.g., 2 mA). **e**, A control policy utilizing a middle state as a noise buffer. Stimulation is high when the control signal is below the bottom threshold and stimulation is low when the control signal is above the top threshold. When the control signal is between the two thresholds, it remains at the level of the stimulation amplitude prior to crossing the threshold (i.e., no changes are made).

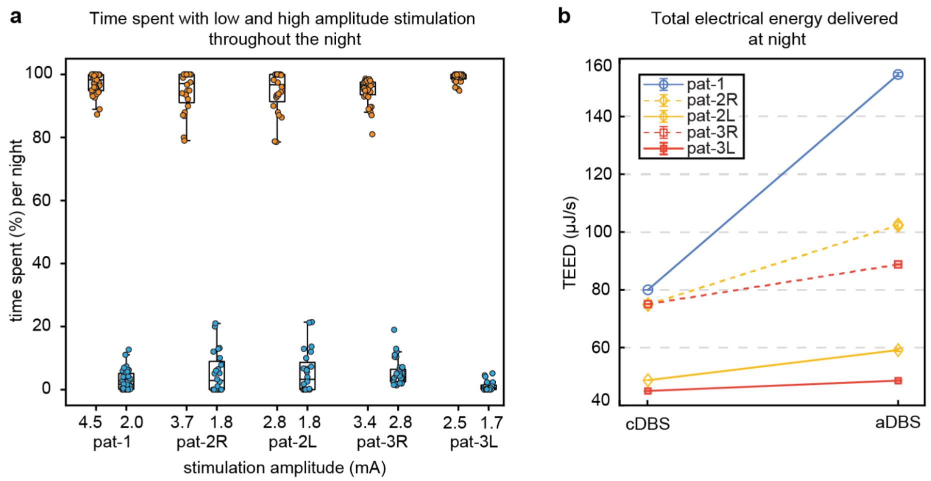


**Extended Data Fig. 7. aDBS algorithm dynamics during the night. a**, Percent time spent at each stimulation amplitude during the night. Each dot represents one night of aDBS testing. Each patient spent a majority of the night in the high stimulation state. **b**, Mean (±standard error of the mean) total electrical energy delivered (TEED) during aDBS and cDBS overnight, showing increased TEED during aDBS in patients, similar to daytime analyses.

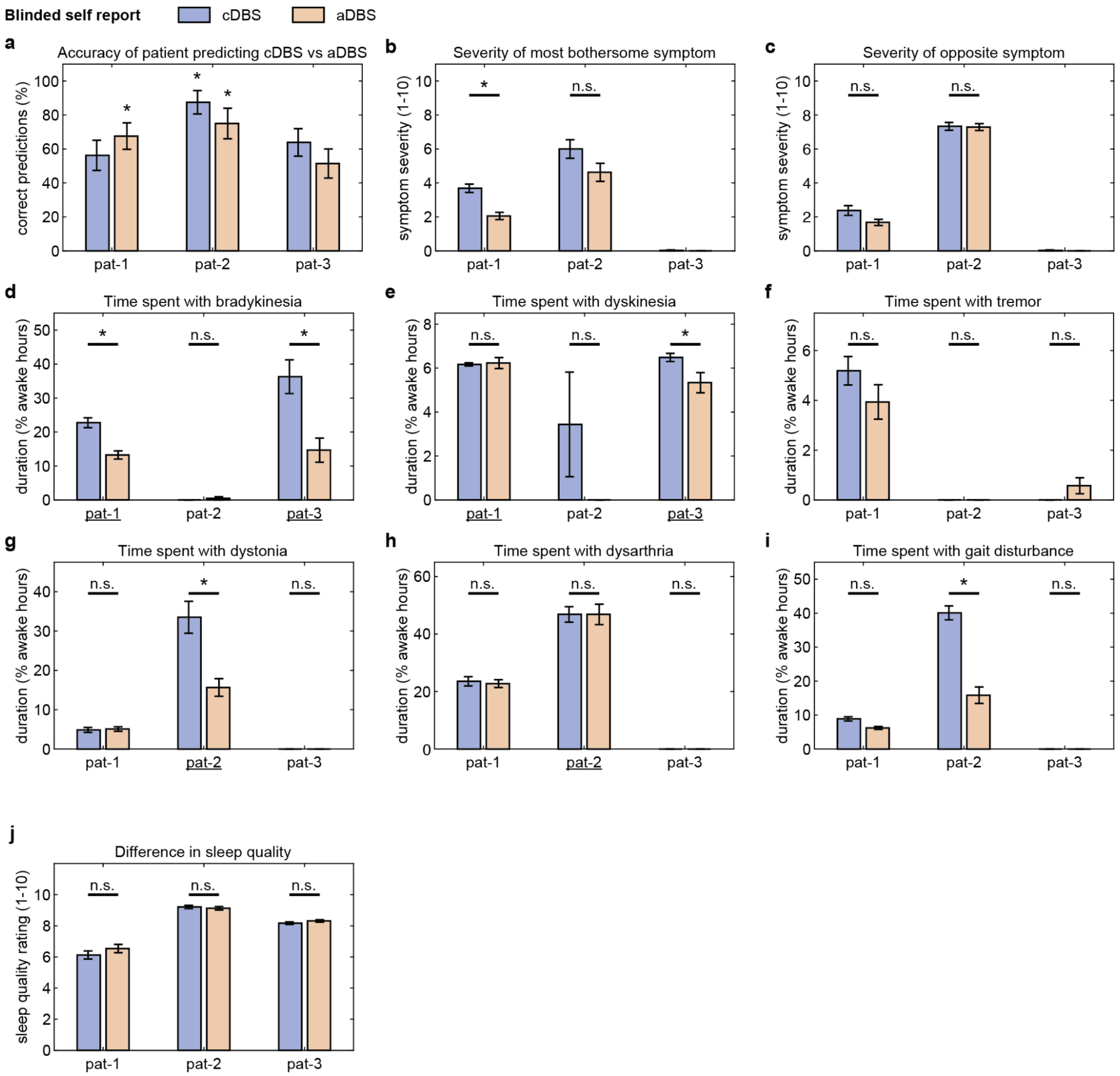


**Extended Data Fig. 8. Effects of aDBS and cDBS on additional motor symptoms and sleep quality. a**, Accuracy of patient inferences of stimulation condition (either aDBS or cDBS). Patient 3 did not correctly conclude stimulation condition at a rate different from chance (cDBS: $p$=0.10, aDBS: $p$=0.87). Patients 1 and 2 correctly inferred aDBS at a rate significantly greater than chance ($p$=0.03 and $p$=0.01, respectively). Additionally, patient 2 correctly deduced cDBS treatment at a rate significantly greater than chance ($p$<0.001). The reason for correctly identifying stimulation condition was never attributed to unusual sensations, but perceived effects on motor symptoms. **b-c**, Patient self-reported motor symptom severity from daily questionnaires (1=least severe, 10=most severe). Patient 3 did not record ratings within the instructed range of 1-10 and data are therefore not reported. **b**, In addition to a decrease in the amount of daily hours with the most bothersome symptom (Fig. 6a), patient 1 also experienced a significant improvement of symptom severity ($p$<0.001). **c**, No subject reported worsened severity of their opposite symptom ($p>0.15$). **d-i**, Comprehensive list of the self-reported duration of symptoms from daily questionnaires. Patients' most bothersome and opposite symptoms are highlighted as underlined patient labels. No symptoms were significantly worsened by aDBS. In addition to improvement in his most bothersome symptom, patient 2 also demonstrated significant improvement in the percentage of waking hours with gait disturbance ($p<0.001$). **j**, Self-reported sleep quality from daily questionnaires (1=poorest sleep, 10=best sleep). aDBS provided no significant change in patients' sleep quality. Error bars reflect standard error of the mean.
